# Establishment & lineage dynamics of the SARS-CoV-2 epidemic in the UK

**DOI:** 10.1101/2020.10.23.20218446

**Authors:** Louis du Plessis, John T. McCrone, Alexander E. Zarebski, Verity Hill, Christopher Ruis, Bernardo Gutierrez, Jayna Raghwani, Jordan Ashworth, Rachel Colquhoun, Thomas R. Connor, Nuno R. Faria, Ben Jackson, Nicholas J. Loman, Áine O’Toole, Samuel M. Nicholls, Kris V. Parag, Emily Scher, Tetyana I. Vasylyeva, Erik M. Volz, Alexander Watts, Isaac I. Bogoch, Kamran Khan, the COVID-19 Genomics UK (COG-UK) Consortium, David M. Aanensen, Moritz U. G. Kraemer, Andrew Rambaut, Oliver G. Pybus

## Abstract

The UK’s COVID-19 epidemic during early 2020 was one of world’s largest and unusually well represented by virus genomic sampling. Here we reveal the fine-scale genetic lineage structure of this epidemic through analysis of 50,887 SARS-CoV-2 genomes, including 26,181 from the UK sampled throughout the country’s first wave of infection. Using large-scale phylogenetic analyses, combined with epidemiological and travel data, we quantify the size, spatio-temporal origins and persistence of genetically-distinct UK transmission lineages. Rapid fluctuations in virus importation rates resulted in >1000 lineages; those introduced prior to national lockdown were larger and more dispersed. Lineage importation and regional lineage diversity declined after lockdown, whilst lineage elimination was size-dependent. We discuss the implications of our genetic perspective on transmission dynamics for COVID-19 epidemiology and control.

## Introduction

Infectious disease epidemics are composed of chains of transmission, yet surprisingly little is known about how co-circulating transmission lineages vary in size, spatial distribution, and persistence, and how key properties such as epidemic size and duration arise from their combined action. Whilst individual-level contact tracing investigations can reconstruct the structure of small-scale transmission clusters (e.g. *1-3*) they cannot be extended practically to large national epidemics. However, recent studies of Ebola, Zika, influenza and other viruses have demonstrated that virus emergence and spread can be instead tracked using large-scale pathogen genome sequencing (e.g. *4-7*). Such studies show that regional epidemics can be highly dynamic at the genetic level, with recurrent importation and extinction of transmission chains within a given location. In addition to measuring genetic diversity, understanding pathogen lineage dynamics can help target interventions effectively (e.g. *8, 9*), track variants with potentially different phenotypes (e.g. *10, 11*), and improve the interpretation of incidence data (e.g. *12, 13*).

The rate and scale of virus genome sequencing worldwide during the COVID-19 pandemic has been unprecedented, with >100,000 SARS-CoV-2 genomes shared online by 1 October 2020 (*14*). Notably, approximately half of these represent UK infections and were generated by the national COVID-19 Genomics UK (COG-UK) consortium (*15*). The UK experienced one of the largest epidemics worldwide during the first half of 2020. Numbers of positive SARS-CoV-2 tests rose in March and peaked in April; by 26 June there had been 40,453 nationally-notified COVID-19 deaths in the UK (deaths occurring ≤28 days of first positive test; *16*). Here, we combine this large genomic data set with epidemiological and travel data to provide a full characterisation of the genetic structure and lineage dynamics of the UK epidemic.

Our study encompasses the initial epidemic wave of COVID-19 in the UK and comprises all SARS-CoV-2 genomes available before 26 June 2020 (50,887 genomes, of which 26,181 were from the UK; Fig 1A). The data represents genomes from 9.29% of confirmed UK COVID-19 cases by 26 June (*16*). Further, using an estimate of the actual size of the UK epidemic (*17*) we infer virus genomes were generated for 0.66% (95% CI=0.46-0.95%) of all UK infections by 5^th^ May.

**Fig. 1.**
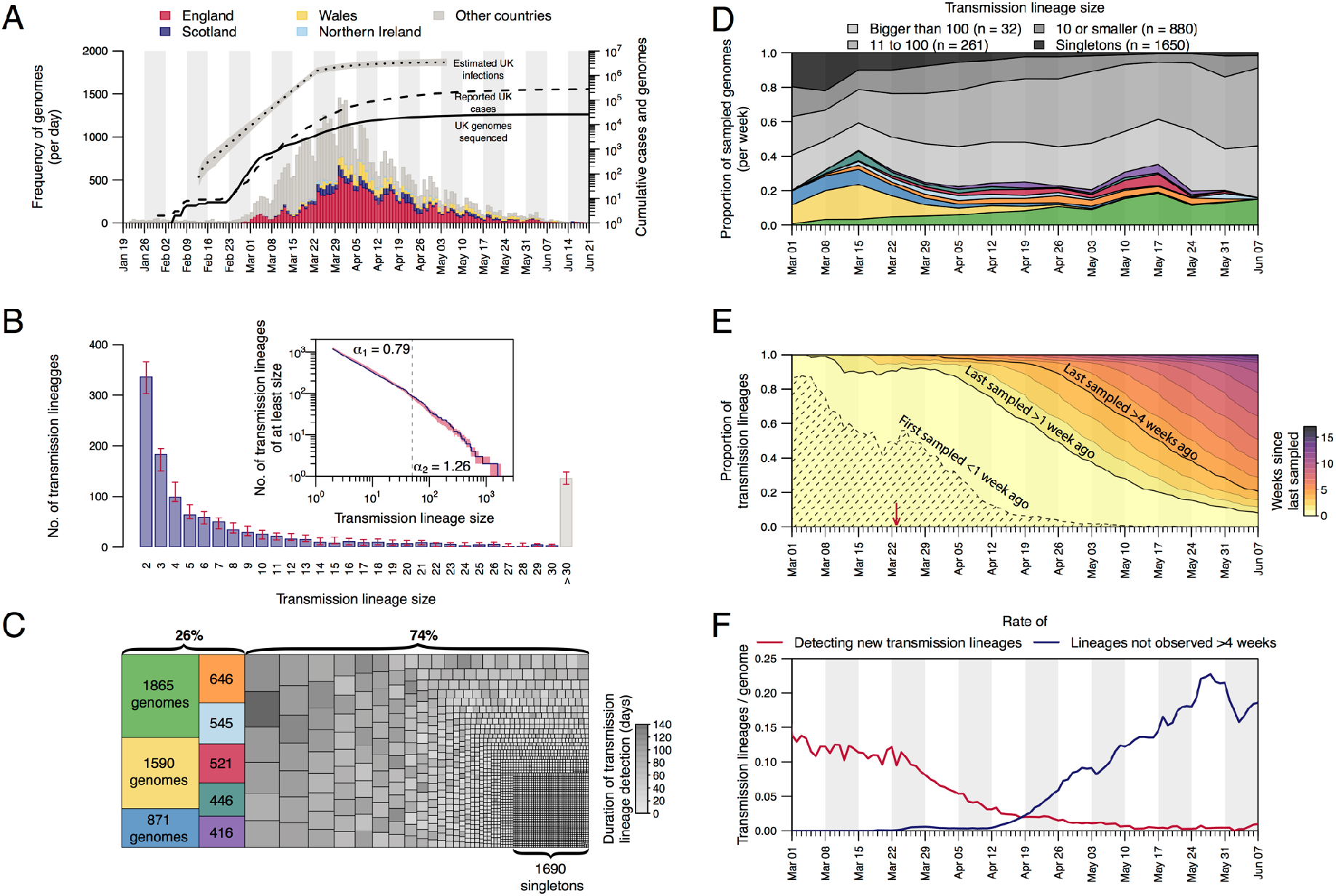
Structure and dynamics of UK transmission lineages. (A) Collection dates of the 50,887 genomes analysed here (left-hand axis). Genomes are coloured by sampling location (England=red, Scotland=dark blue, Wales=yellow, Northern Ireland=light blue, elsewhere=grey). The solid line shows the cumulative number of UK virus genomes (right-hand axis). The dashed and dotted lines show, respectively, the cumulative number of laboratoryconfirmed UK cases (by specimen date) and the estimated number of UK infections (17; grey shading=95% CI; right-hand axis). Due to retrospective screening, the cumulative number of genomes early in the epidemic exceeds that of confirmed cases. (B) Distribution of UK transmission lineage sizes. Blue bars show the number of transmission lineages of each size (red bars=95% HPD of these sizes across the posterior tree distribution). Inset: the corresponding cumulative frequency distribution of lineage size (blue line), on double logarithmic axes (red shading=95% HPD of this distribution across the posterior tree distribution). Values either side of vertical dashed line show coefficients of power-law distributions (P[X ≥ x] ∼ x^α^) fitted to lineages containing ≤50 (α_1_) and >50 (α_2_) virus genomes, respectively. (C) Partition of 26,181 UK genomes into UK transmission lineages and singletons, coloured by (i) lineage, for the 8 largest lineages, or (ii) duration of lineage detection (time between the lineage’s oldest and most recent genomes) for the remainder. (D) Lineage size breakdown of UK genomes collected each week. Colours of the 8 largest lineages are as depicted in (C). (E) Trends through time in the detection of UK transmission lineages. For each day, all lineages detected up to that day are coloured by the time since the transmission lineage was last sampled. Isoclines correspond to weeks. Shaded area=transmission lineages that were first sampled <1 week ago. The red arrow indicates the start of the UK lockdown. (F) Red line=daily rate of detecting new transmission lineages. Blue line=rate at which lineages have not been observed for >4 weeks.

### Genetic structure and lineage dynamics of the UK epidemic

We first sought to identify and enumerate all independently introduced, genetically-distinct chains of infection within the UK. We developed a large-scale molecular clock phylogenetic pipeline to identify “UK transmission lineages” that (i) contain two or more UK genomes and (ii) descend from an ancestral lineage inferred to exist outside of the UK (Fig. S1, S2). Sources of statistical uncertainty in lineage assignation were taken into account. We identified a total of 1179 (95% HPD=1143-1286) UK transmission lineages. Although each is intended to capture a chain of local transmission arising from a single importation event, some UK transmission lineages will be unobserved or aggregated due to limited SARS-CoV-2 genetic diversity (*18*) or incomplete or uneven genome sampling (*19, 20*). Therefore we expect this number to be an underestimate (see Methods). In our phylogenetic analysis 1650 (95% HPD=1611-1783) UK genomes could not be allocated to a UK transmission lineage (singletons).

Most transmission lineages are small and 72.4% (95% HPD=69.3-72.9%) contain <10 genomes (Fig. 1B). However the lineage size distribution is strongly skewed and follows a power-law distribution (Fig. 1B inset), such that the 8 largest UK transmission lineages contain >25% of all sampled UK genomes (Fig. 1C, Figs. S4-S7 show further visualisations). Although the two largest transmission lineages are estimated to comprise >1500 UK genomes each, there is phylogenetic uncertainty in their sizes (95% HPDs=1280-2133 and 1342-2011 genomes). All 8 largest lineages were first detected before the UK national lockdown on 23 March and, as expected, larger lineages were observed for longer (Pearson’s r=0.82; 95% CI=0.8-0.83; Fig. S9). The sampling frequency of lineages of varying sizes differed over time (Fig. 1D); whilst UK transmission lineages containing >100 genomes consistently accounted for >40% of weekly sampled genomes, the proportion of small transmission lineages (≤10 genomes) and singletons decreased over the course of the epidemic (Fig. 1D).

The detection of UK transmission lineages in our data changed markedly through time. In early March the epidemic was characterised by lineages first observed within the previous week (Fig. 1E). The per-genome rate of appearance of new lineages was initially high, then declined throughout March and April (Fig. 1F), such that by 1^st^ May 96.2% of sampled genomes belonged to transmission lineages that were first observed >7 days previously. By 1^st^ June, a growing number of lineages (>73%) had not been detected by genomic sampling for >4 weeks, suggesting that they were rare or had gone extinct, a result that is robust to the sampling rate (Fig. 1F, 1A). Together, these results indicate that the UK’s first epidemic wave resulted from the concurrent growth of many hundreds of independently-introduced transmission lineages, and that the introduction of non-pharmaceutical interventions (NPIs) was followed by the apparent extinction of lineages in a size-dependent manner.

### Transmission lineage diversity and geographic range

We also characterised the spatial distribution of UK transmission lineages using available data on 107 virus genome sampling locations, which correspond broadly to UK counties or metropolitan regions. Although genomes were not collected randomly (some lineages and regions will be over-represented due to targeted investigation of local outbreaks; e.g. *21*) the number of UK lineages detected in each region correlates with the number of genomes sequenced (Fig. 2A, Pearson’s r=0.96, 95% CI=0.95-0.98) and the number of reported cases (Fig. S10, Pearson’s r=0.6, 95% CI=0.44-0.72) in each region. Further, larger lineages were observed in more locations; every 100 additional genomes in a lineage increases its observed range by 6-7 regions (Fig. 2B; Pearson’s r=0.8, 95% CI=0.78-0.82). Thus, bigger regional epidemics comprised a greater diversity of transmission lineages, and larger lineages were more geographically widespread. These observations indicate substantial dissemination of a subset of lineages across the UK and suggest many regions experienced a series of introductions of new lineages from elsewhere, potentially hindering the impact of local interventions.

**Fig. 2.**
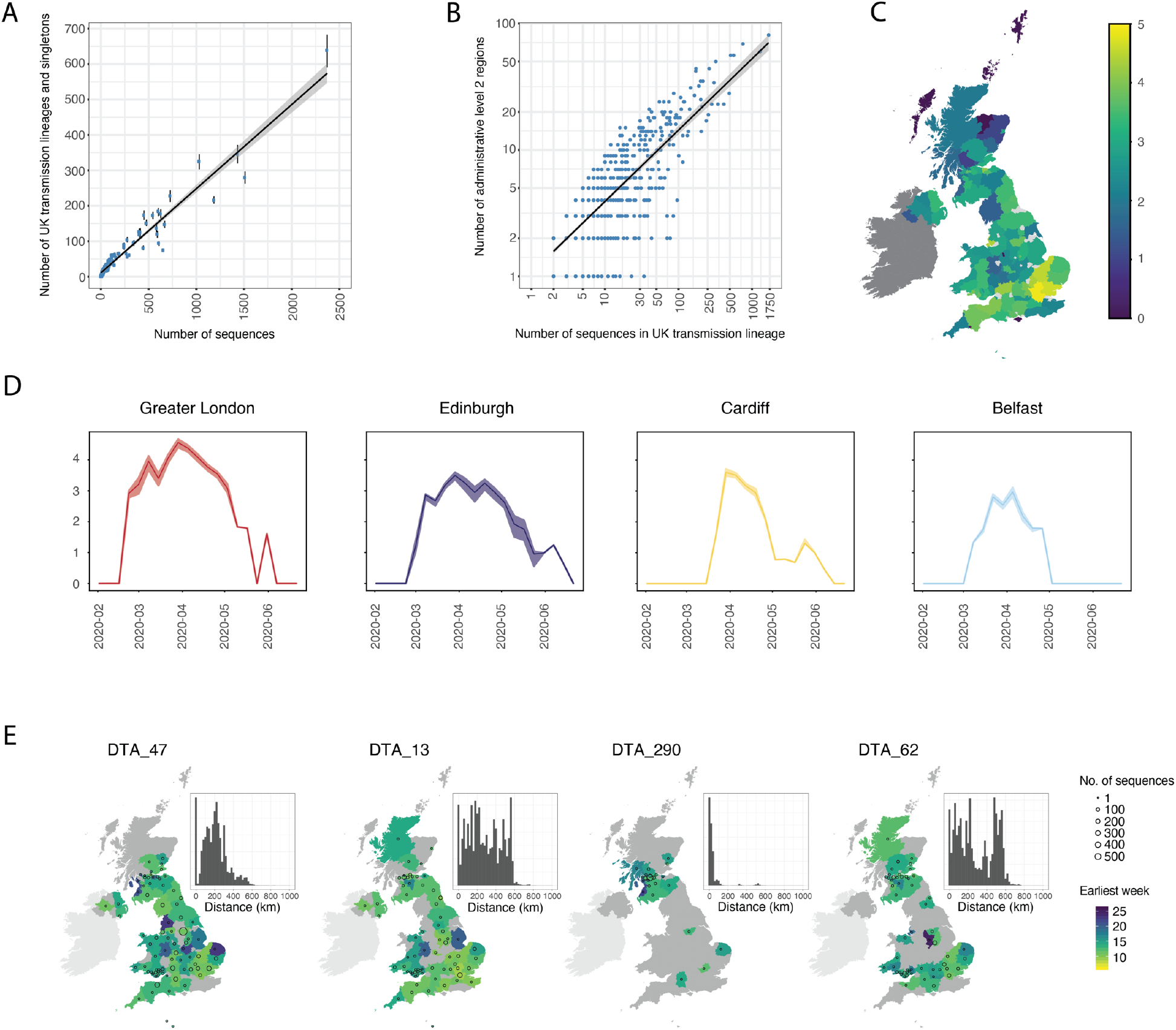
Spatial distribution of UK transmission lineages. (A) Correlation between the number of transmission lineages detected in each region (points=median values, bars=95% HPD intervals) and the number of UK virus genomes from each region (Pearson’s r=0.96, 95% CI=0.95-0.98, p<0.001). (B) Correlation between the spatial range of each transmission lineage and the number of virus genomes it contains (Pearson’s r=0.8, 95% CI=0.78-0.82, p<0.001) (C) Map showing Shannon’s index (SI) for each region, calculated across the study period (2^nd^ Feb-26^th^ Jun). Yellow colours indicate higher SI values and darker colours lower values. (D) SI through time for the UK national capital cities. (E) Illustration of the diverse spatial range distributions of UK transmission lineages. Colours represent the week of the first detected genome in the transmission lineage in each location. Circles show the number of sampled genomes per location. Insets show the distribution of geographic distances for all sequence pairs within the lineage (see Fig S12 for further details).

We quantified the substantial variation among regions in the diversity of transmission lineages present using Shannon’s index (SI; this value increases as both the number of lineages and the evenness of their frequencies increase; Fig. 2C). We observed the highest SIs in Hertfordshire (4.77), Greater London (4.62) and Essex (4.49); these locations are characterised by frequent commuter travel to/within London and proximity to major international airports (*22*). Locations with the three lowest non-zero SIs were in Scotland (Stirling=0.96, Aberdeenshire=1.04, Inverclyde=1.32; Fig. 2C).

To illustrate temporal trends in transmission lineage diversity, we plot SI through time for each of the UK’s national capital cities (Fig. 2D). Lineage diversities in each peaked in late March and declined after the UK national lockdown, congruent with Figure 1E, F. Greater London’s epidemic was the most diverse and characterised by an early, rapid rise in SI (Fig. 2C), consistent with epidemiological trends there (*16, 23*). Belfast’s lineage diversity was notably lower.

We observe variation in the spatial range of individual UK transmission lineages. Although some lineages are widespread, most are more localised and the range size distribution is right-skewed (Fig. S11), congruent with an observed abundance of small lineages (Fig. 1B, 2B) and biogeographic theory (e.g. *24*). For example, lineage DTA_13 is geographically dispersed (>50% of sequence pairs sampled >234km apart) whereas DTA_290 is strongly local (95% of sequence pairs sampled <100km apart) and DTA_62 has multiple foci of sampled genomes (Fig. 2E, S12). The national distribution of cases therefore arose from the aggregation of multiple heterogeneous lineage-specific patterns.

### Dynamics of international introduction of transmission lineages

The process by which transmission lineages are introduced to an area is an important aspect of early epidemic growth (e.g. *25*). To investigate this at a national scale we estimated the rate and source of SARS-CoV-2 importations into the UK. Since standard phylogeographic approaches were precluded by strong biases in genome sampling among countries (*19*), we developed a new approach that combines virus phylogenetics with epidemiological and travel data. First, we estimated the TMRCA (time to the most recent common ancestor) of each UK transmission lineage. The TMRCAs of most UK lineages are dated to March and early April (median=21^st^ March; IQR=14^th^-29^th^ March). UK lineages with earlier TMRCAs are larger and longer-lived than those whose TMRCAs postdate the national lockdown (Fig. 3A, S15).

**Fig. 3.**
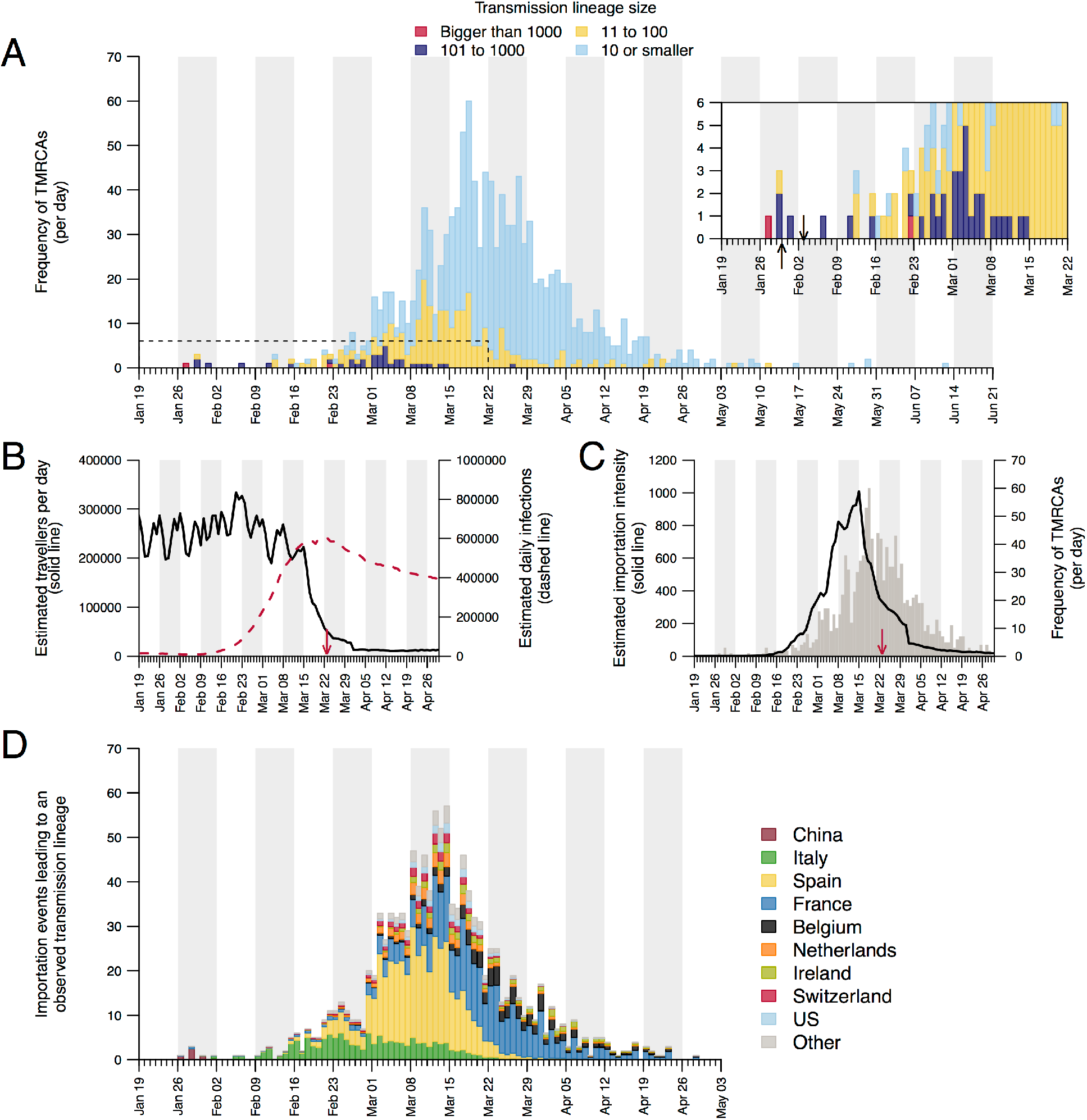
Dynamics of transmission lineage importation (A) Histogram of lineage TMRCAs, coloured by lineage size. Inset: expanded view of the days prior to UK lockdown. Left-hand arrow=collection date of the UK’s first laboratory-confirmed case; right-hand arrow=collection date of the earliest UK virus genome in our dataset. (B) Estimated number of inbound travellers to the UK per day (black) and estimated number of infectious cases worldwide (dashed red). Arrow here shows the start of the UK lockdown. (C) Estimated importation intensity (EII) curve (black) and the histogram of lineage TMRCAs (grey). (D) Estimated histogram of virus lineage importation events per day, obtained from our lag model. Colours show the proportion attributable each day to inbound travel from various countries (see Figs. S19, S20, Table S4). This assignment is statistical, i.e. we cannot ascribe a specific source location to any given lineage.

Due to incomplete sampling, TMRCAs best represent the date of the first inferred transmission event in a lineage, not its importation date (Fig. S2). To infer the latter, and quantify the delay between importation and onward within-UK transmission, we generated daily estimates of the number of travellers arriving in the UK and of global SARS-CoV-2 infections (see Methods) worldwide. Before March, the UK received ∼1.75m inbound travellers per week (school holidays explain the end-February ∼10% increase; Fig. 3B). International arrivals fell by ∼95% during March and this reduction was maintained through April. Elsewhere, estimated numbers of infectious cases peaked in late March (Fig. 3B). We combined these two trends to generate an estimated importation intensity (EII) - a daily empirical measure of the intensity of SARS-CoV-2 importation into the UK. The EII peaks in mid-March, when high UK inbound travel volumes coincided with growing numbers of infectious cases elsewhere (Fig. 3B, C).

Crucially, the EII’s temporal profile closely matches, but precedes, that of the TMRCAs of UK transmission lineages (Fig. 3A, C). The difference between the two represents the “importation lag”, the time elapsed between lineage importation and the first detected local transmission event (Fig. S2). Using a statistical model, we estimate importation lag to be on average 8.22 ± 5.21 days (IQR=3.35-15.18) across all transmission lineages. Further, importation lag is strongly size-dependent; average lag is ∼10 days for lineages comprising ≤10 genomes and <1 day for lineages of >100 genomes (Table S2). This size-dependency likely arises because the earliest transmission event in a lineage is more likely to be captured if it contains many genomes (Fig. S2; see Methods). We use this model to impute an importation date for each UK transmission lineage (Fig. 3D). Importation was unexpectedly dynamic, rising and falling substantially over only 4 weeks, hence 80% of importations (that gave rise to detectable UK transmission lineages) occurred between 27 February and 30 March. The delay between the inferred date of importation and the first genomic detection of each lineage was 14.13 ± 5.61 days on average (IQR=10-18) and declined through time (Table S2, S3).

To investigate country-specific contributions to virus importation we generated separate importation intensity (EII) curves for each country (Fig. S17). Using these values, we estimated the numbers of inferred importations each day attributable to inbound travel from each source location. As with the rate of importation (Fig. 3A), the relative contributions of arrivals from different countries were dynamic (Fig. 3D). Dominant source locations shifted rapidly in February and March and the diversity of source locations increased in mid-March (Fig. S17). Earliest importations were most likely from China or elsewhere in Asia but were rare compared to those from Europe. Over our study period we infer ∼33% of UK transmission lineages stemmed from arrivals from Spain, 29% from France, 12% from Italy and 26% from elsewhere (Fig. S20; Table S4). These large-scale trends were not apparent from individual-level travel histories; routine collection of such data ceased on 12 March (*26*).

## Conclusions

The exceptional size of our genomic survey provides insight into the micro-epidemiological patterns that underlie the features of a large, national COVID-19 epidemic, allowing us to quantify the abundance, size distribution, and spatial range of transmission lineages. Prelockdown, high travel volumes and few restrictions on international arrivals (Table S5; Fig. 3B) led to the establishment and co-circulation of >1000 identifiable UK transmission lineages (Fig. 3A), jointly contributing to accelerated epidemic growth that quickly exceeded national contact tracing capacity (*26*). The relative contributions of importation and local transmission to initial epidemic dynamics under such circumstances warrants further investigation. We expect similar trends occurred in other countries with comparably large epidemics and high international travel volumes; virus genomic studies from regions with smaller or controlled COVID-19 epidemics have reported high importation rates followed by more transient lineage persistence (e.g. *27*-*29*).

Earlier lineages were larger, more dispersed, and harder to eliminate, highlighting the importance of rapid or pre-emptive interventions in reducing transmission (e.g. *30-32*). The high heterogeneity in SARS-CoV-2 transmission at the individual level (*33*-*35*) appears to extend to whole transmission lineages, such that >75% of sampled viruses belong to the top 20% of lineages ranked by size. Whilst the national lockdown coincided with limited importation and reduced regional lineage diversity, its impact on lineage extinction was size-dependent (Fig. 1E, F). The over-dispersed nature of SARS-CoV-2 transmission likely exacerbated this effect (*36*), thereby favouring, as *R*_*t*_ declined, greater survival of larger widespread lineages and faster local elimination of lineages in low prevalence regions. The degree to which the surviving lineages contributed to the UK’s ongoing second epidemic is currently under investigation. The transmission structure and dynamics measured here provide a new context in which future public health actions at regional, national, and international scales should be planned and evaluated.

## Supporting information

Full list of names and affiliations of COG-UK consortium members.

GISAID acknowledgments table

## Data Availability

UK SARS-CoV-2 genomes and public metadata are available from www.cogconsortium.uk/data/ and deposited at gisaid.org and ENA (bioproject PRJEB37886). Non-UK genomes were obtained from gisaid.org. Raw data and analysis files for this work are in supplementary materials or available from GitHub at https://github.com/COG-UK/UK-lineage-dynamics-analysis, which also contains a list of sequence accession numbers. A GISAID acknowledgments table is available at https://github.com/COG-UK/UK-lineage-dynamics-analysis

https://gisaid.org

https://github.com/COG-UK/UK-lineage-dynamics-analysis

## Acknowledgments

We are grateful to everyone worldwide involved in generating the virus genome data shared on GISAID. We thank Samir Bhatt, Philippe Lemey, and Christopher Dye for insightful discussion. Funding: COG-UK is funded by the Medical Research Council (MRC) part of UK Research & Innovation (UKRI), the National Institute of Health Research (NIHR) and Genome Research Limited, operating as the Wellcome Sanger Institute. VH was supported by BBSRC grant BB/M010996/1; CR by a Fondation Botnar Research Award (Programme grant 6063) and UK Cystic Fibrosis Trust (Innovation Hub Award 001); JR by the UKRI GCRF One Health Poultry Hub (BB/S011269/1); MUGK by a Branco Weiss Fellowship and EU H2020 project MOOD; NRF by WT fellowship 204311/Z/16/Z and MRC-FAPESP awards MR/S0195/1 and 18/14389-0; TIV by a Branco Weiss Fellowship; IIB is funded by the Canadian Institutes for Health Research (02179 – 000); JTM, RMC, NJL and AR by WT Collaborators Award 206298/Z/17/Z; AR and ES by ERC grant 725422; DMA by NIHR Global Health Research Unit (16/136/111); OGP, MUGK, LDP and AEZ by the Oxford Martin School. Author contributions: Study design: LdP, JTM, MUGK, AR, OGP. Methods development/programming: LdP, JTM, MUGK, AR, OGP, AEZ, VH, CR, JA, RC, TC, BJ, NJL, AO, SN, DMA, ES. Data analysis: LdP, MUGK, AR, OGP, AEZ, VH, CR, DMA, JTM, BG, KVP, ES, TIV. Data collection/experiments: AW, IIB, KK. Wrote paper: LdP, JR, MUGK, OGP. Edited paper/figure creation: LdP, MUGK, JR, CR, BG, TIV, NRF, EMV. Competing interests: KK is the founder of BlueDot, a social enterprise that develops digital technologies for public health. AW and IIB received employment or consulting income from BlueDot during this research. Data and materials availability: UK SARS-CoV-2 genomes and public metadata are available from www.cogconsortium.uk/data/ and deposited at gisaid.org and ENA (bioproject PRJEB37886). Non-UK genomes were obtained from gisaid.org. Raw data and analysis files for this work are in supplementary materials or available from GitHub at https://github.com/COG-UK/UK-lineage-dynamics-analysis, which also contains a list of sequence accession numbers.

## Methods

### Genomic data

All SARS-CoV-2 genomes available on GISAID (*14*) on 23 June 2020 were downloaded and combined with all SARS-CoV-2 genomes sequenced by the COG-UK consortium (*15*) by 26 June 2020 (available at https://www.cogconsortium.uk/data/). The pipeline used to collect and process raw SARS-CoV-2 sequence data and sample-associated metadata across the national COG-UK network is described in (*37*). We removed sequences that were from duplicate or environmental samples, those without exact collection dates, and those with large clusters of substitutions or large indels. Each genome sequence was aligned to the reference (Wuhan-Hu-1, GenBank: MN908947.3) using *minimap v2.17* (*38*) and the resulting SAM alignment was converted to a FASTA alignment, with the 5’ and 3’ UTRs of each genome masked by Ns. Insertions relative to the reference were discarded and site 11,083 (site position relative to MN908947), which is globally homoplasic, was also masked. Genomes that contained >5% Ns after mapping and those with a genetic distance to WH04 (GISAID: EPI_ISL_406801) more than 4 standard deviations from the epi-week mean genetic distance to WH04 were discarded. The final dataset consisted of 50,887 genomes sampled between 24 December 2019 and 22 June 2020, of which 26,181 (∼51%) were from the UK (see Fig. 1A).

### Geographical metadata

Administrative level 2 (admin2) metadata for the sampling location of UK virus genome sequences in the dataset (roughly equivalent to counties in the UK) required cleaning in order to be mapped to official admin2 regions, as found in the Global Administrative Database (GADM, https://gadm.org).

Some sampling locations in the metadata could not be unambiguously mapped to a known location (e.g. “City Centre”), while others were for locations in overseas territories (e.g. Falklands and Gibraltar). Yet other genome sequences had uninformative spatial records (e.g. Yorkshire or Wales), or no admin2 level data at all. For these (3431 of 26,181) the admin2 region was not mapped. We carried out a simple one-to-one mapping where possible, which included correcting spelling mistakes and alternative entries for the same county (e.g. Durham versus County Durham). Locations recorded at a higher spatial resolution were mapped to the corresponding admin2 region (e.g. Solihull was mapped to Birmingham). Where the recorded locations were larger than the admin2 regions (e.g. “West Midlands”), and most of the sequences in the area were from this larger conglomeration as opposed to its higher-resolution components, these admin2 regions were combined. When creating the map figures, we also merged some city authorities with no reported sequences with their surrounding county, on the assumption that the larger county was used to represent the location of city samples (e.g. for Leicester and Leicestershire). Finally, genome sequences from Northern Ireland reported locations as historical counties, rather than the official admin2 designations, and so these historical counties were used instead. The cleaning code is provided on the GitHub repository (https://github.com/COG-UK/UK-lineage-dynamics-analysis).

### Phylogenetic analysis and molecular clock dating

We developed a new Bayesian molecular clock phylogenetic analysis pipeline in order to reconstruct a posterior set of time-scaled phylogenetic trees for our exceptionally large virus genome dataset. Using the standard Bayesian approach it is currently impractical to estimate time-scaled trees directly from genome sequence data for more than a few thousand sequences. Therefore, we employed a number of extensions to make the analysis tractable.

First, we divided the full genome sequence dataset (n = 50,887) into five smaller datasets. Genomes were assigned SARS-CoV-2 lineages according to the nomenclature defined in *(39)* using *Pangolin* (*40;* github.com/covlineages/pangolin). Each lineage (and its sublineages) represents a monophyletic clade in the global SARS-CoV-2 phylogeny and can thus be analysed independently. For each lineage in A (n = 3591), B (n = 8821), B.1 (n = 22,861), B.1.1 (n = 15,616), we estimated an approximately maximum-likelihood tree using the Jukes-Cantor model in *FastTree v2.1.10* (*41*), then collapsed branch lengths shorter than 5×10^−6^ substitutions per site, which corresponded to distances smaller than one substitution across the whole virus genome, and likely result from nucleotide ambiguity codes in the genome sequences. By pruning out a large monophyletic clade the maximumlikelihood tree for B.1 was further divided into two trees, B.1.pruned (n = 12,275) and B.1.X (n = 10,586).

Prior to analysing the full dataset, an initial analysis was performed on a subset of genomes to obtain estimates of the molecular clock rate and of the TMRCA of each large-scale phylogenetic tree defined above. The full dataset was subsampled as evenly as possible across epi-weeks and countries with a slight enrichment for samples immediately descended from five large polytomies in the global phylogeny. For each of these nodes, we always included the five oldest genomes, the most recent genome sequence and five other immediate descendants that were randomly chosen. The remaining genomes were sampled by allocating an even number of sequences per epi-week while maintaining a dataset size of <1,000 genomes. For each epi-week, genomes were sampled evenly by country until either its allocation was exhausted or there were no remaining genomes available. This subsampled dataset was analysed in *BEAST 1.10* (*42*) using a GTR+G+F substitution model, with a strict molecular clock model using a non-informative continuous-time Markov chain (CTMC) prior (*43*) and a Skygrid coalescent tree prior (*44*) with 40 grid points, roughly corresponding to weeks between 1 October 2019 and 2 July 2020. In the analysis, monophyly constraints were used to ensure that the clades corresponding to the large-scale phylogenetic trees identified in the previous step were monophyletic. We combined four independent Markov Chain Monte Carlo (MCMC) chains that were each run for 40 million steps, discarding the first 4 million steps of each chain as burn-in and resampling states every 4000 steps. Convergence was assessed using *Tracer* (*45*).

Next, we applied a commonly used approach, recently implemented in *BEAST 1.10*, to convert branches of the large-scale phylogenetic trees from units of substitutions per site to time. This model takes the place of the nucleotide substitution model in a traditional Bayesian molecular clock dating analysis. Briefly, each branch of a maximum-likelihood tree is first scaled to represent the number of substitutions that occurred along that branch. Polytomies are resolved by inserting branches of length 0 substitutions. The likelihood of a branch *b*_*i*_ of length *s*_*i*_ substitutions is defined by a Poisson distribution with mean *t*_*i*_*m* where *t*_*i*_ is the length of the branch in years and *m* is the clock rate. The log-likelihood of the whole tree is then the sum of the log-likelihoods of each branch, which represents a fixed, strict-clock model and follows a commonly implemented approach for scaling phylogenies into time-calibrated trees (e.g. *46-48*).

Each large-scale phylogenetic tree was analysed under a strict clock model, with the clock rate fixed to the median estimate from the preliminary analysis (7.5×10^−4^ substitutions/site/year) and a Laplace root-height prior with mean equal to the median TMRCA estimate of the corresponding subtree in the preliminary analysis and scale equal to the average distance from the median. Trees were sampled using MCMC under the model described above with a Skygrid coalescent tree prior (*44*) using the same grid-points as in the preliminary analysis. A randomly resolved time-calibrated tree estimated in *TreeTime* (*49*) was used as the starting tree. To maintain a mapping between the topology in the estimated time-calibrated tree and the input genetic distance tree, we constrained the topologies such that any tree-move that broke a clade present in the input tree was rejected. The resulting MCMC chain, therefore, only samples different polytomy resolutions and branch durations. This approach allowed us to incorporate uncertainty in the polytomy resolutions and branch durations into our molecular clock analysis.

We ran between 8 and 24 chains for 60 to 100 million MCMC steps for each large-scale phylogeny. Upon completion, we discarded 15 million states as burn-in from each chain. Chains that did not converge or pass the burn-in in less than 15 million states were re-run. Chains were combined and resampled every 100,000 states using custom R-scripts, leaving between 6808 and 17,020 posterior samples of each large-scale phylogenetic tree. Convergence was assessed using *Tracer* (*45*) and the R-package *coda* (*50*).

### Identifying transmission lineages

We define a “UK transmission lineage” as two or more UK infection cases that (i) descend from a shared, single importation of the virus into the UK from elsewhere, (ii) are the result of subsequent local transmission within the UK, and (iii) were present in our virus genome sequence dataset. This concept is illustrated in Figure S1 and is distinct from a transmission cluster, an epidemiological term commonly referring to a group of cases that occur close to each other in space and time (e.g. in a hospital or care home). Therefore, a large UK transmission lineage may comprise many different individual transmission clusters.

[It is important to note that the “UK transmission lineage” definition employed here is distinct from the lineage/phylotype designations used by other parts of the COG-UK consortium and that are displayed at https://microreact.org/project/cogconsortium. Those latter designations (which have the format “UK…”) are defined on the basis of shared sets of mutations, rather than shared descent from an inferred single introduction event.]

We can identify UK transmission lineages in the time-calibrated trees estimated in the previous step as clades of two or more genomes sampled in the UK. The TMRCA of all genome sequences in a UK transmission lineage represents the earliest transmission event in the lineage revealed by the data; however, it does not necessarily represent the first transmission event in the lineage as a whole, nor does it represent the importation date (i.e. the arrival date of the index patient in the UK). The relationship between the TMRCA of a UK transmission lineage in our dataset and the importation date is illustrated in Figure S2. Specifically, if the transmission lineage is well-sampled, then the TMRCA represents the date of the first transmission event in the lineage (TMRCA A in Fig. S2, UK transmission lineage 2 in Fig. S1). However, if the transmission lineage is sparsely sampled then the TMRCA may represent a later transmission event (TMRCA B in Fig. S2, UK transmission lineage 1 in Fig. S1). The “importation date” of each UK transmission lineage is the date that an infected inbound traveller entered the UK.

We used a two-state asymmetric discrete trait analysis (DTA) model (*51*) implemented in *BEAST 1.10* (*42*) to infer ancestral node locations (UK, non-UK) on empirical distributions of 500 time-calibrated trees sampled from each of the posterior tree distributions estimated above. Additionally, we used a robust counting approach (*52*) to estimate the expected number of location state transitions into and out of the UK. For each large-scale subtree, we combined 2 independent chains, each run for 5 million MCMC steps and sampled every 4500 states. The first 10% of each run was discarded as burn-in, resulting in 2000 trees with estimates of the ancestral location for each internal node. Finally, *TreeAnnotator 1.10* was used to generate maximum clade credibility (MCC) trees for each subtree, where each internal node is assigned a posterior probability of representing a transmission event in the UK.

Transmission lineages were identified by first labelling each node in the MCC trees as UK or non-UK and then initiating a depth-first search from each UK genome in the MCC trees. All nodes with a median age after 23 January 2020 and posterior probability >0.5 of the ancestral location being located in the UK were labelled as UK nodes. The depth-first search is continued until a non-UK node is encountered or there are no nodes left to explore. At the end of the depth-first search, all nodes visited by the search are added to the same (arbitrarily named) UK transmission lineage. If only one tip is visited, the UK genome at the tip is marked as a singleton. This procedure is repeated iteratively until every UK genome in the tree has been assigned a transmission lineage or marked as a singleton. The same procedure was repeated on each of the 2000 posterior trees, for each subtree, from the DTA analyses described above to examine statistical uncertainty in the number, size and duration of UK transmission lineages and their TMRCA distribution.

Our methodology is likely to underestimate the true number of transmission lineages and singletons. Since only a small fraction of UK infections have been sequenced (Fig. 1A), many lineages will have gone undetected. Furthermore, the power to detect a transmission lineage in our sparsely-sampled dataset is dependent on its size (i.e. the frequency of a lineage being sampled from a small random sample of infections), making it more likely for larger lineages to be detected. The low sampling fraction means that some singletons detected in our dataset likely belong to observed and unobserved UK transmission lineages. Nonetheless, the true number of singletons (importations not resulting in onward transmission) is likely to be significantly more than our estimate, because their small size makes them difficult to detect with a low sampling fraction. Finally, under-sampling of genomes from other countries could result in mistaken aggregation of separate importations, reducing the number of detected lineages. This mistaken aggregation will result in larger, older lineages being estimated. This was the motivation for placing an age limit on UK nodes in the tree. We chose 23 January 2020 as the oldest possible date for a transmission event in the UK as this represents the date that the first patient who tested positive for SARS-CoV-2 in the UK entered the country (*53*) (tested positive on 30 January 2020). Although older importations into the UK could in theory be possible, if they had resulted in large autochthonous outbreaks, we would have observed this in both epidemiological and genomic data.

We estimate a median of 2968 (95% HPD 2829-3103) non-UK to UK state transitions and an additional 1468 (95% HPD 1362-1566) UK to non-UK state transitions (Fig. S3, Table S1) using the robust counting approach (*52*). The former slightly exceeds the sum of transmission lineages and singletons as identified on the MCC tree (=2918) and across the 2000 posterior trees (median=2829, 95% HPD=2773-3048; Table S1). This result is expected, since multiple location state changes along long branches contribute to the total number of state transitions, but do not add to the total number of UK transmission lineages or singletons. The largest number of location state transitions occur on the B.1.1 phylogeny, with the fewest occurring on lineage A, which are the largest and smallest of the subtrees, respectively. Proportional to the number of tips, fewer state changes are inferred on the two B.1 phylogenies than other subtrees, while the number of UK to non-UK transitions on the B phylogeny exceeds that inferred on other lineages. We caution that UK to non-UK transitions are likely to be underestimated because of under-sampling in other countries and differences in the proportion of infections sequenced between countries.

The transmission lineage size distribution from the MCC trees falls within the HPD interval taken across the 2000 posterior trees (Fig. 1B). Although the sizes of the largest transmission lineages vary substantially across posterior trees, the cumulative size distributions are similar across all trees (Fig. 1B, inset). Similarly, the transmission lineage duration distribution on the MCC trees falls within the variation of the HPD interval taken across the 2000 posterior trees (Fig. S8).

We used the Jaccard index to compare the classification of UK genome sequences into transmission lineages and singletons between posterior trees and the MCC trees. Figure S13A shows the mean, median and 95% HPD interval of the Jaccard index for each posterior tree compared to the 1999 other posterior trees, across all subtrees. While most Jaccard indices are between 0.7 and 0.8, there is a noteworthy minority of trees with mean Jaccard indices <0.6 (n=100). Comparing the 2000 posterior trees to the classification on the MCC tree (Fig. S13B), results in a similar distribution of Jaccard indices, with most indices between 0.7 and 0.8 and minorities below 0.6 and above 0.8 (n=68, n=170 respectively).

We undertook a similar analysis of the sensitivity to phylogenetic uncertainty of the distribution of UK transmission lineage TMRCAs. We computed the median and 95% HPD interval of the number of transmission lineage TMRCAs on each date across the 2000 sampled posterior trees. Figure S14 shows that the TMRCA distribution computed from the MCC trees falls within the comparatively narrow HPD limits, and oscillates around the median estimate for each date.

### UK epidemiological data

The number of reported COVID-19 cases in the UK, by specimen date, were downloaded from https://coronavirus.data.gov.uk/cases (date accessed: 1 September 2020). The number of reported COVID-19 cases for each Upper Tier Local Authority (UTLA) in England, Local Health Board (LHB) in Wales and regional NHS Board in Scotland, by specimen date, were downloaded from https://coronavirus.data.gov.uk/downloads/csv/coronavirus-cases_latest.csv, http://www2.nphs.wales.nhs.uk:8080/CommunitySurveillanceDocs.nsf (file: *“Rapid COVID-19 surveillance data.xlsx”*) and https://github.com/DataScienceScotland/COVID-19-Management-Information (file: *“COVID19 - Daily Management Information - Scottish Health Boards - Cumulative cases.csv”*), respectively (date accessed: 15 October 2020).

To enable comparison of case and sequence data, locations used to report case data were combined to correspond to those used for sequence data and vice-versa (see the *Geographical metadata* section). Northern Ireland was not included due to inconsistencies between the locations used for case and sequence data reporting that could not be easily resolved.

### Global deaths due to COVID-19

The cumulative number of daily COVID-19 deaths for each country were downloaded from the JHU CSSE COVID19 Database (date accessed: 19 August 2020) (*54*). We removed data pertaining to cruise ships, and aggregated data to the country level where data were reported for subnational divisions (e.g. Australia). For countries with overseas territories included in the dataset (e.g. United Kingdom), we excluded the cumulative death counts in those overseas territories. For each country we computed a time series of the daily number of deaths by taking the difference in the cumulative number on consecutive days. When this difference was negative, for example when corrections in the cumulative number were not propagated backwards, we set the value to zero. A relevant outlier in these time series is the addition of 1290 deaths in China on 17 April 2020, while on the days before and after no deaths were recorded. To account for these deaths, we uniformly distributed these deaths over the previous 85 days described by the epidemiological data.

### Population data

Country population size estimates were downloaded from the UN Department of Economic and Social Affairs website (https://population.un.org/wpp/Download/Standard/Population/), using the *Medium* fertility projection for 2020 (*55*).

### Travel and mobility data

To investigate temporal trends in SARS-CoV-2 importation intensity we sought information on the number of travellers entering the UK from each other country for the period from 1 January to 30 April 2020. Incoming travellers comprised both British nationals and resident and visiting citizens of other countries. Estimates were obtained by combining multiple data sources. First, the UK Home Office has provided statistics that describe the number of inbound travellers arriving in the UK by air on each day during this period (https://assets.publishing.service.gov.uk/government/uploads/system/uploads/attachment_data/file/887655/statisticsrelating-to-covid-19-and-the-immigration-system-tables-may-2020-arrivals.ods). This data set provides the daily number of incoming air passengers but not their source country. Second, we obtained the number of tickets sold for inbound flight journeys to the UK along with their origin location from the IATA (for passengers that transfer, the source location is the country from where the whole journey started). We used these numbers to calculate the percentage of arrivals from each country on a monthly basis from January to April 2020. We multiplied the monthly distribution of source destination by the total number of air passenger arrivals in the UK each day to estimate the number of arrivals from each country. Third, we augmented the above air passenger numbers with estimated numbers of incoming travellers arriving per day by short-sea ferry and through the Channel Tunnel (French: *Le tunnel sous la Manche*). Numbers of short-sea ferry passengers from France, Netherlands and the Republic of Ireland were estimated from monthly statistics obtained from the UK Department of Transport (https://assets.publishing.service.gov.uk/government/uploads/system/uploads/attachment_data/file/908445/spas0107.ods). Within that data set, values are provided for the Republic of Ireland and for “Other EU countries”. The latter total was broken down by country using data from 2019 showing that 72.7% of UK short-sea journeys are with France, 13.6% with the Republic of Ireland, 10.4% with the Netherlands, and 3.3% with other countries (https://www.gov.uk/government/statistics/sea-passenger-statistics-2019-short-sea-routes). Eurotunnel Shuttle vehicle movements from France were obtained from publicly available monthly records (https://www.eurotunnelfreight.com/uk/2020/02/shuttle-traffic-for-january-2020). In the absence of other information we assumed (i) inbound and outbound vehicle movements via the Eurotunnel Shuttle services were equally frequent and (ii) one passenger per truck and 1.5 passengers per passenger vehicle. Inbound Eurostar rail passenger numbers from France and Belgium were estimated from available data and adjusted as far as possible for post-pandemic reduction in travel. Specifically, ∼2m passengers travelled by Eurostar in the first quarter of 2020 (https://www.breakingtravelnews.com/news/article/eurostar-passenger-count-slips-by-a-fifth-in-early-2020).

Monthly Eurostar passenger numbers were then calculated by assuming (i) inbound and outbound journeys were equally frequent, (ii) two thirds of inbound Eurostar journeys originated in France and one third in Belgium, in approximate proportion to the ratio of services, and (iii) the proportional decrease in Eurostar travel volumes during March and April 2020 was equal to that observed for vehicle movements via the Eurotunnel Shuttle. Our estimates do not incorporate estimates of movements across the land border between the UK and the Republic of Ireland. This is unlikely to be problematic as the numbers of infections in the Republic of Ireland was relatively low compared to other potential source countries during the time period of interest.

### Epidemiological model

We sought an estimate of the number of individuals in each source country who are (i) infected with SARS-CoV-2 and (ii) able to travel to the UK and initiate a transmission chain. In what follows we refer to these individuals as the “potential initiators of a transmission lineage” (PITL). We conservatively assumed that symptomatic individuals cannot initiate a transmission chain in the UK, either through being prevented from travelling or perfect isolation on arrival. Thus, our estimates of daily SARS-CoV-2 prevalence includes only pre-symptomatic and asymptomatic individuals. Asymptomatic individuals are counted among the PITL as those capable of initiating a transmission lineage at any time while they are still infectious. Figure S16 illustrates the ways in which individuals are counted towards the daily PITL and their potential disease outcomes.

We estimated the daily number of PITL by back-extrapolating the time series of daily numbers of deaths due to COVID-19 in each source country. COVID-19 deaths were used instead of confirmed cases, as we are primarily interested in temporal dynamics rather than absolute values, and death counts are believed to be less sensitive to changes in case definition, reporting delays and differences in the level of surveillance among countries and regions. Estimates of the latent period (infection to becoming infectious), incubation period (infection to onset of symptoms), the infectious duration, and the time between symptom onset and death (in fatal cases) were used to estimate the number of infected individuals who would go on to die from COVID-19, in each stage of the disease, on each day (Fig. S16). We then estimated the total number of infected individuals on each day by multiplying with the reciprocal of the infection fatality rate (IFR).

Estimates of the periods defined above were taken from peer-reviewed sources. Specifically, we assumed that the time from acquiring an infection to becoming infectious is 3 days (*56*) and the time to symptom onset 5 days (2 days after becoming infectious) (*57*). The infectious period for patients who recover from the disease was assumed to end 5 days after symptom onset (*56*) while those who die from the disease are assumed to do so 18 days after the onset of symptoms (*58*). Given the large numbers of deaths we expect that variation in these timings among individuals will be averaged out and is not considered. We further assumed an asymptomatic proportion of 31% (*59*) and an IFR of 1%, which is broadly consistent with those found in the literature for China, France, and passengers aboard the Diamond Princess (*58, 60, 61*). These values correspond to our study period, the spring epidemic of COVID-19; more recent estimates of IFR may vary due to changing treatment regimes and other factors. To examine the sensitivity of our results to the asymptomatic proportion we re-ran our analysis with proportions of 0.18 and 0.78 (the range of published estimates; *62, 63*), and found that our results were robust over this range (data not shown). As our main results are fully determined by temporal trends in EII and not absolute numbers, they are invariant to the value of the IFR and we did not perform a sensitivity analysis on it. We did not account for changing levels of infectivity among individuals over the course of their infection.

Using the time series of deaths extracted from the JHU CSSE COVID-19 Database (*54*), as described above, we obtained estimates of the daily number of PITL in 183 countries from 31 December 2019 to 26 July 2020.

### Estimated importation intensity

The daily “estimated importation intensity” (EII) of a country is defined as the product of the proportion of individuals in that country who make up the PITL (as described above) on each day, and the number of individuals who travelled from that country to the UK on that day. The former is estimated by dividing our estimate of the total number of individuals who could potentially initiate a lineage (for each day) by the total population of the country (see the *Epidemiological model* section). The latter corresponds to the total number of arrivals by air, ferry, and rail on that day (see the *Travel and mobility data* section). To assist in the subsequent use of the EII, we aggregated all countries with low PITL estimates into a single “other” category. The aggregated countries are those that comprised less than 1% of the cumulative total number of cases as of 1 May 2020 (excluding the UK). This left 53 primary source locations. Maximum EII (**Fig. S17**) was highest for Spain, (which experienced a large, early epidemic that peaked before inbound passenger numbers declined), followed by France (whose later epidemic peak coincided with high but declining international travel).

### Importation lag model

We modelled the TMRCA of an observed transmission lineage (the data observation) as the arrival date of the index patient (of that transmission cluster) in the UK, *G*, plus a lag time, *L*, until the first transmission event in the lineage revealed by the data. Given the probability that an importation occurs on day *g, f*_*G*_(*g*), and the probability of a lag time of *j* days, *f*_*L*_(*j*), the probability of a TMRCA occurring on day *k* is *v*_*k*_, is defined by

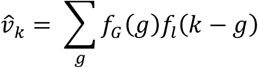

with 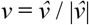. TMRCAs and importation dates are assumed to be independent, so the likelihood for all transmission lineages is the product of the corresponding *v*_*k*_ for each lineage.

This model does not account for incomplete sampling of patients from UK transmission lineages. It is likely that the TMRCA of a small transmission lineage is more recent than the first transmission event after the importation and this issue is potentially further exacerbated by non-random sampling of genome sequences from patients in the lineage (*64*). We therefore expect shorter lag times for bigger transmission lineages. To account for this size-dependence, we model the average importation lag as a function of lineage size. The functional form of this is given by the equation *α* + *β* / *n*, where *α* corresponds to the minimal average lag time expected under complete sampling of the lineage and *β* accounts for the increase in lag time as a smaller proportion of sequences are included in the lineage.

We applied this model to the TMRCA estimates of individual transmission lineages and their sizes as obtained from the MCC trees (see the *Identifying transmission lineages* section). Values for *α* and *β* were found by numerically optimising the likelihood function using random draws from an exponential distribution as initial parameter values. The optimisation procedure was repeated several times to ensure that the algorithm did not become stuck in a local optimum. We further tested whether lineage size affects the importation lag through a likelihood ratio test (LRT) comparing the above model to a nested model without size dependence (*β* = 0) and found that the size-dependent model is preferred (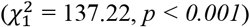, *p < 0.001*). The maximum likelihood estimates for *α* and *β* are 0.72 and 28.91 (Fig. S18), respectively.

### Travel advice in the UK

The travel advice issued by the Foreign and Commonwealth Office (FCO) of the United Kingdom pertaining to countries and regions affected by COVID-19 was primarily made available through their website (*FCO Travel advice: coronavirus (COVID-19)* at https://www.gov.uk/guidance/travel-advice-novel-coronavirus). The number of COVID-19 cases in the UK was available via the government website (*Coronavirus cases in the UK* at https://www.gov.uk/guidance/coronavirus-covid-19-information-for-the-public). Travel advice was also echoed by various news outlets and other information platforms, such as the Public Health Scotland/NHS Scotland *Fit for Travel* website (https://www.fitfortravel.nhs.uk/). We collected this information by mining archived FCO sites, manually retrieving HTML files corresponding to updates to the URLs provided above and available at the Internet Archive (https://archive.org/). Files were obtained and examined for all dates when changes to the URL were published (18 updates were published in total between 4 February and 23 May 2020). Furthermore, we compared this advice with the *Fit for Travel* online resource, collected through a similar approach. Where information was insufficient or unclear, we complemented it with data from news outlets to clarify travel advice, which was the case before February 4, when there was no official travel advice (only notifications for novel coronavirus). We collated all the travel advice information into a single standardised table containing types of advice, dates of implementation and countries or geographic regions covered by the advice. The types of advice included both suggestions against specific types of travel versus all but non-essential travel and the recommended period of self-isolation upon return from specific destinations. All of the changes in travel advice were between February 6 and March 23, when specific self-isolation recommendations applied to the general population and not just returning travellers. A summary of the main changes in the UK travel advice across time (in particular, dates when advice for new countries were issued) is presented in Table S5.

---

**Fig. S1.**
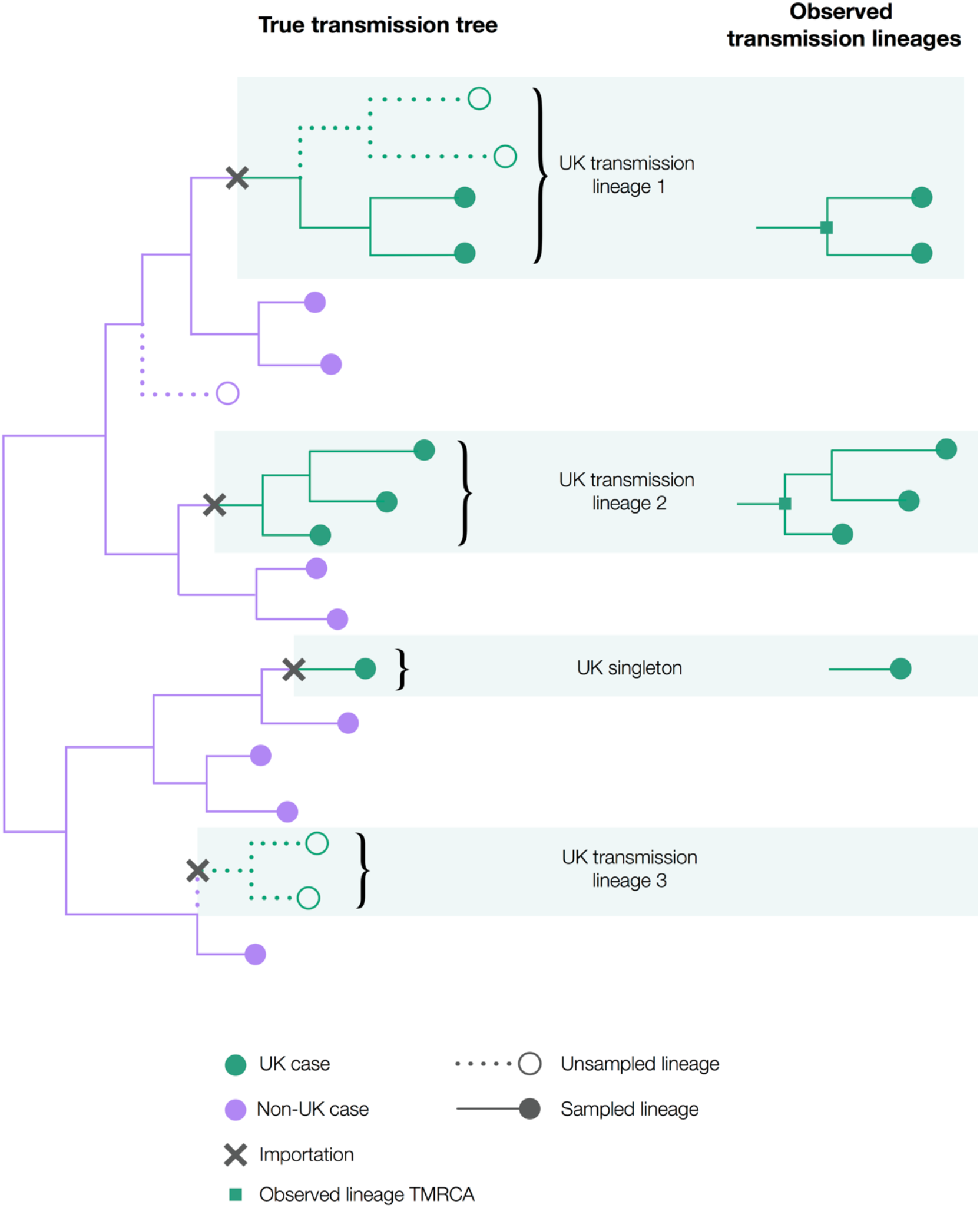
Hypothetical scenario illustrating the definition and international context of UK transmission lineages. Note that only half of cases in UK transmission lineage 1 are observed and that UK transmission lineage 3 is not observed at all. Singletons do not contribute to onward transmission within the UK and are not classified here as UK transmission lineages.

**Fig. S2.**
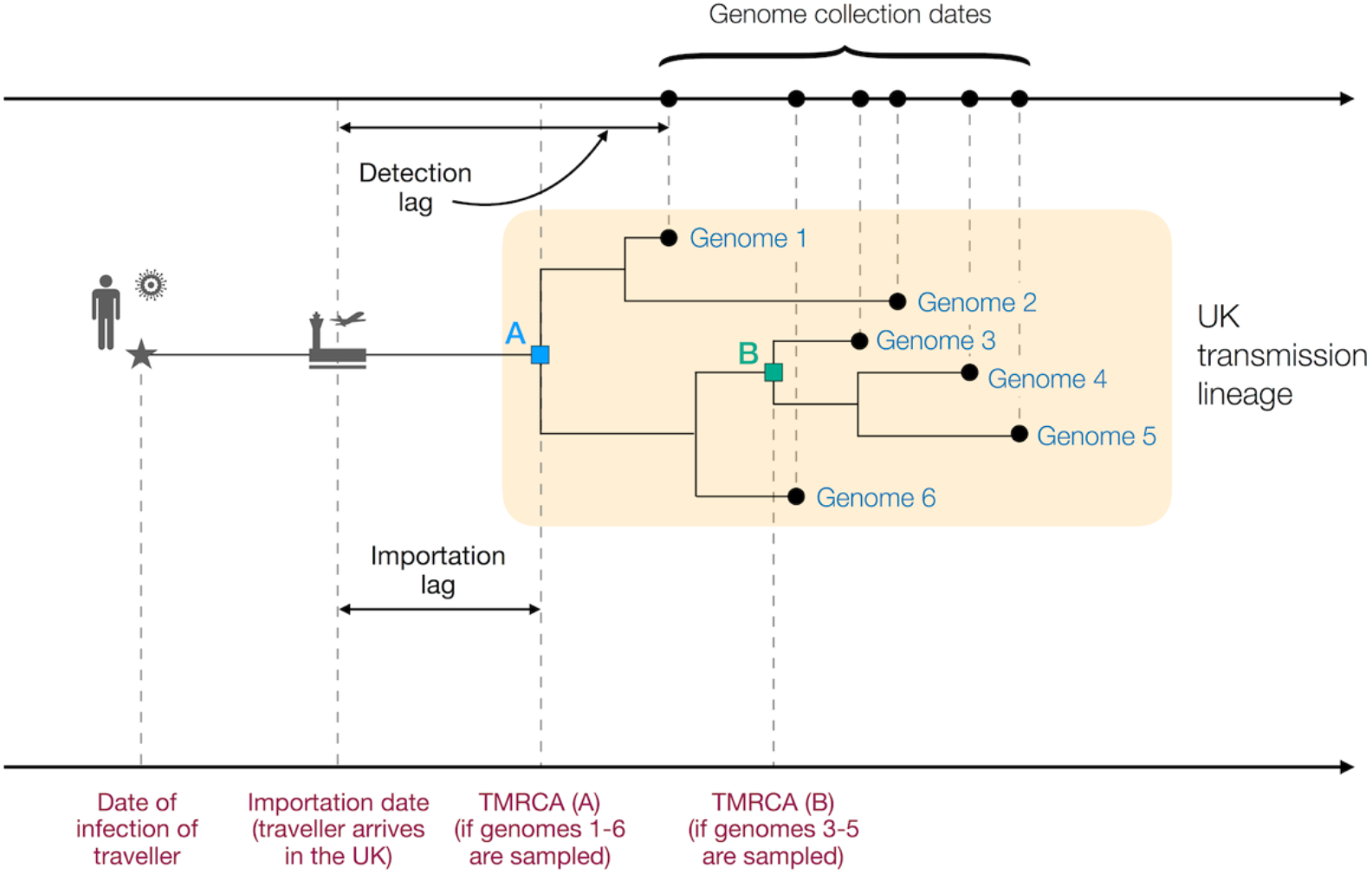
Figurative illustration of a UK transmission lineage detected through genome sampling. To be detected, a UK transmission lineage must contain two or more sampled genomes (see **Figure S1**). The terms TMRCA, detection lag, and importation lag can be understood with reference to this figure. The lineage TMRCA is sample dependent, for example, TMRCA A is observed if genomes 1–6 are sampled and TMRCA B is observed if only genomes 3–5 are sampled.

**Fig. S3.**
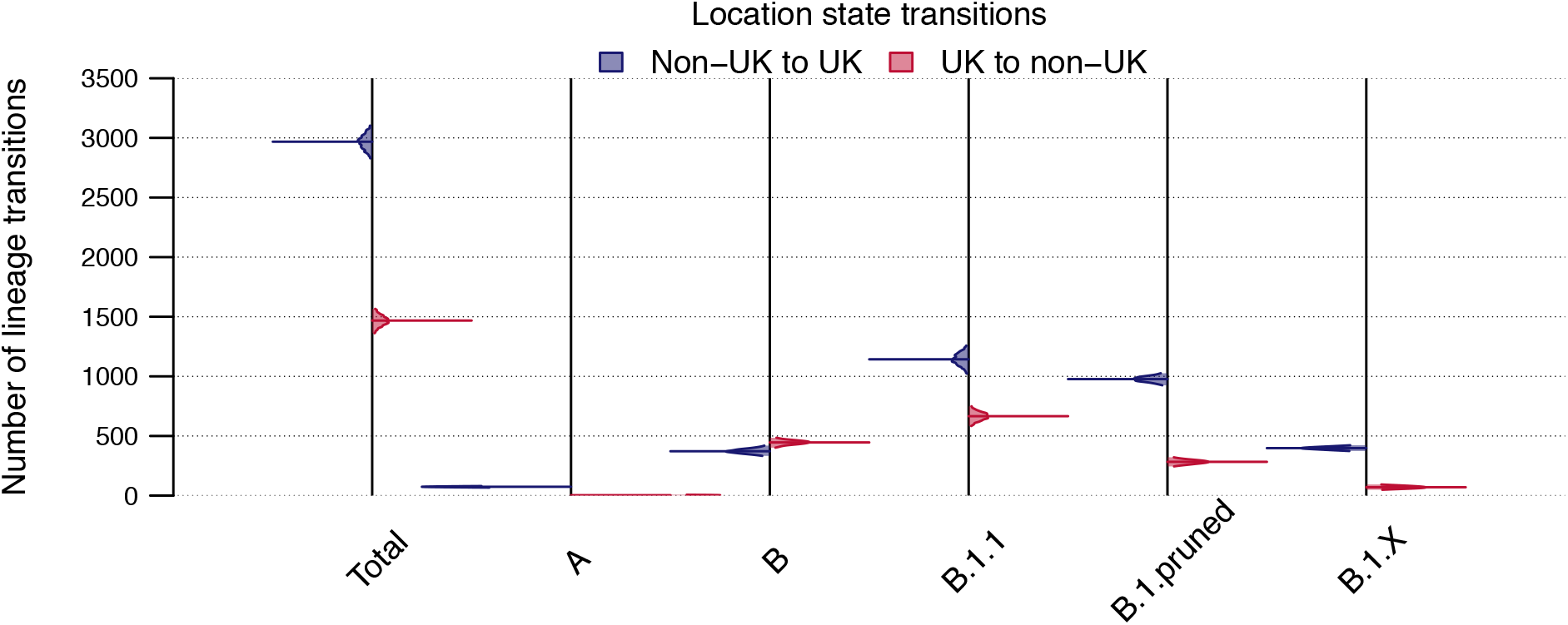
Number of location state transitions between the binary phylogenetic traits UK/non-UK detected by the robust counting approach implemented in BEAST 1.10. Non-UK to UK=blue, UK to non-UK=red. Posterior distributions are truncated at their 95% HPD interval limits and the horizontal lines indicate median estimates.

**Fig. S4.**
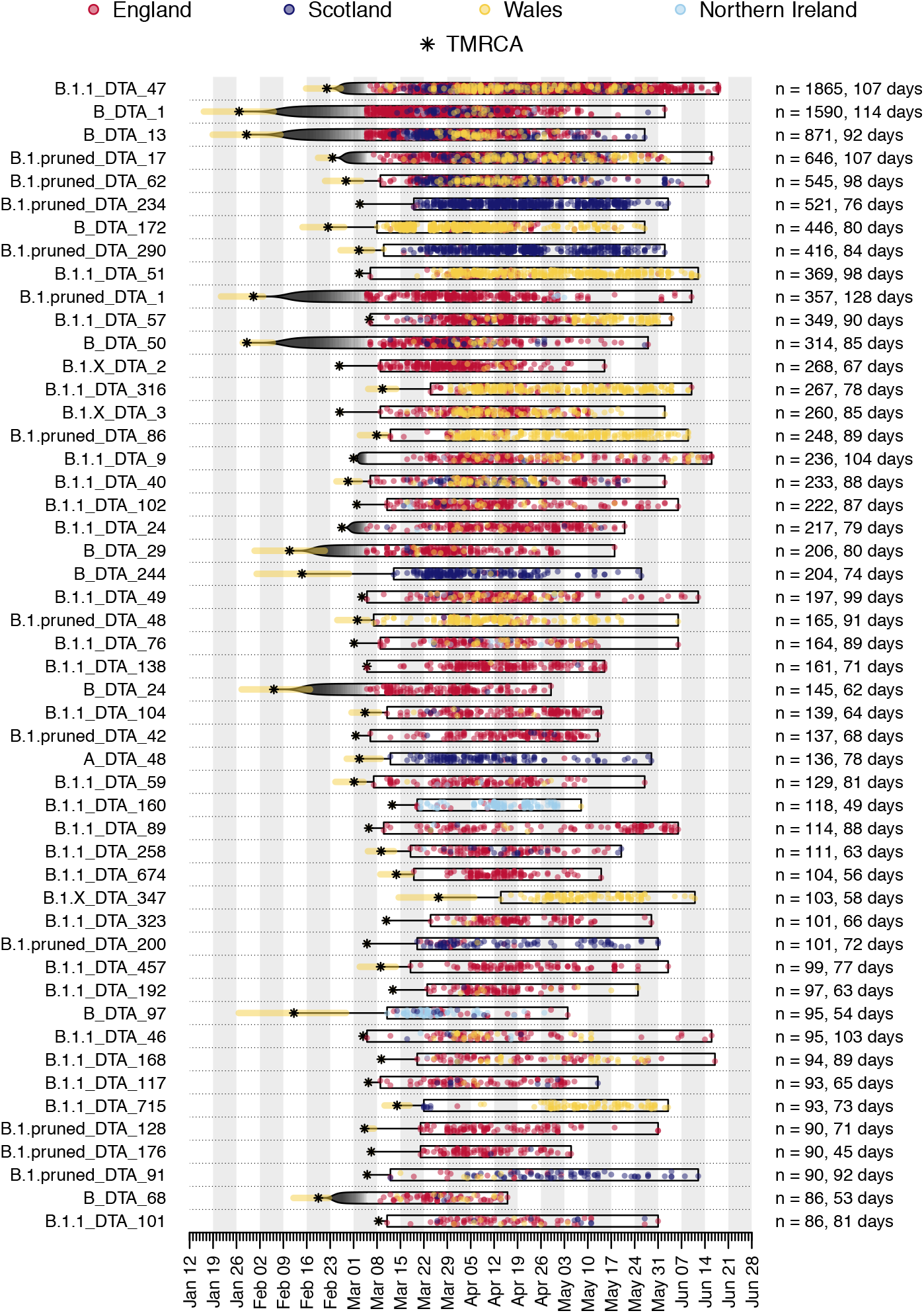
Illustration of the time course of the 50 largest UK transmission lineages in our dataset. Each row is a transmission lineage. Dots are genome sampling times (coloured by sampling location) and boxes show the range of sampling times for each transmission lineage (sampling duration). Asterisks show the median TMRCA of each lineage and the yellow bars show the 95% HPD of each TMRCA. On the right, n indicates the number of UK genomes in the lineage and the duration of lineage detection (time between the lineage’s oldest and most recent genomes). Sampling times of the first 500 SARS-CoV-2 genomes collected in the UK have been obscured.

**Fig. S5.**
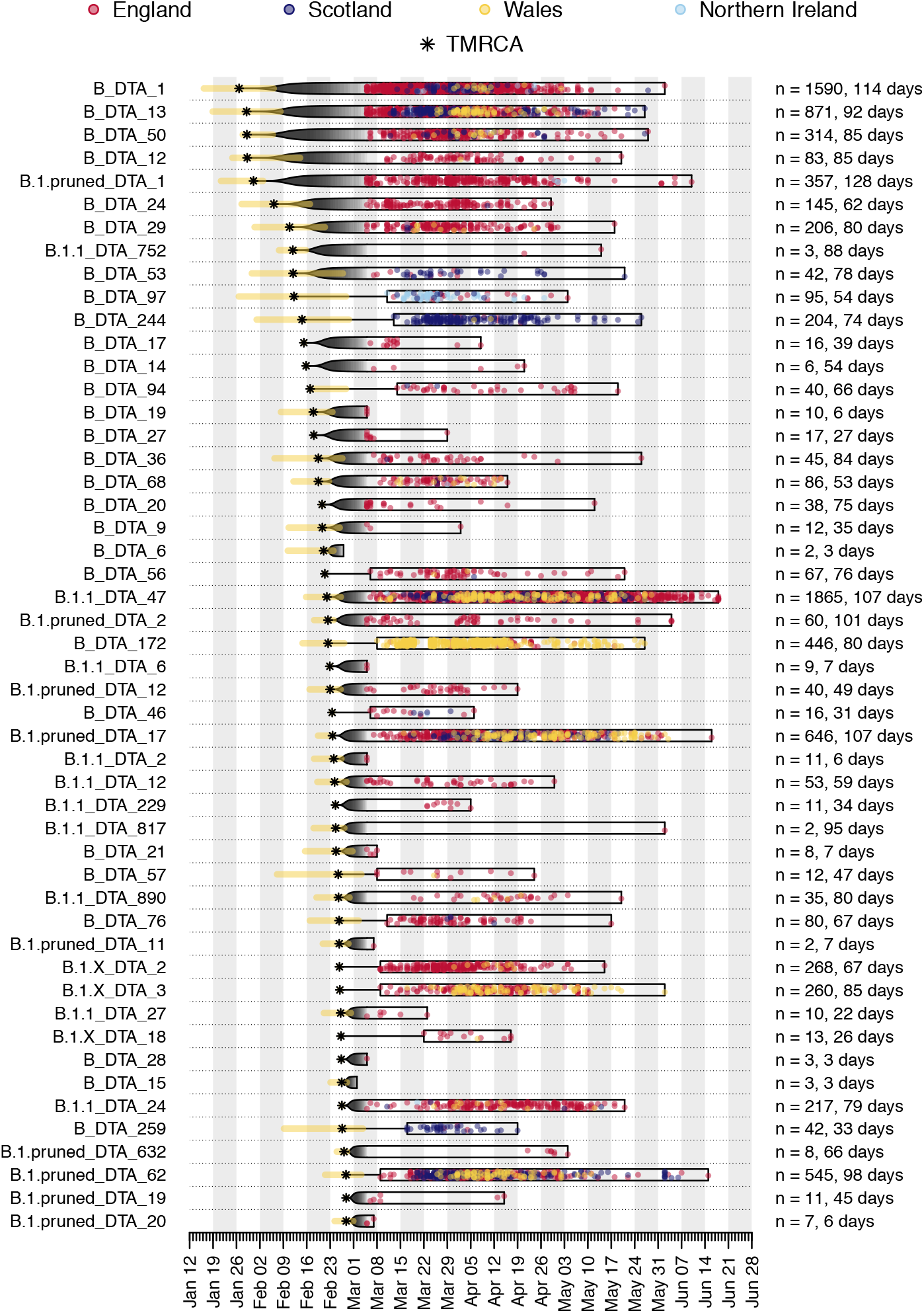
Illustration of the time course of the 50 earliest UK transmission lineages in our dataset. See Figure S4 caption for details.

**Fig. S6.**
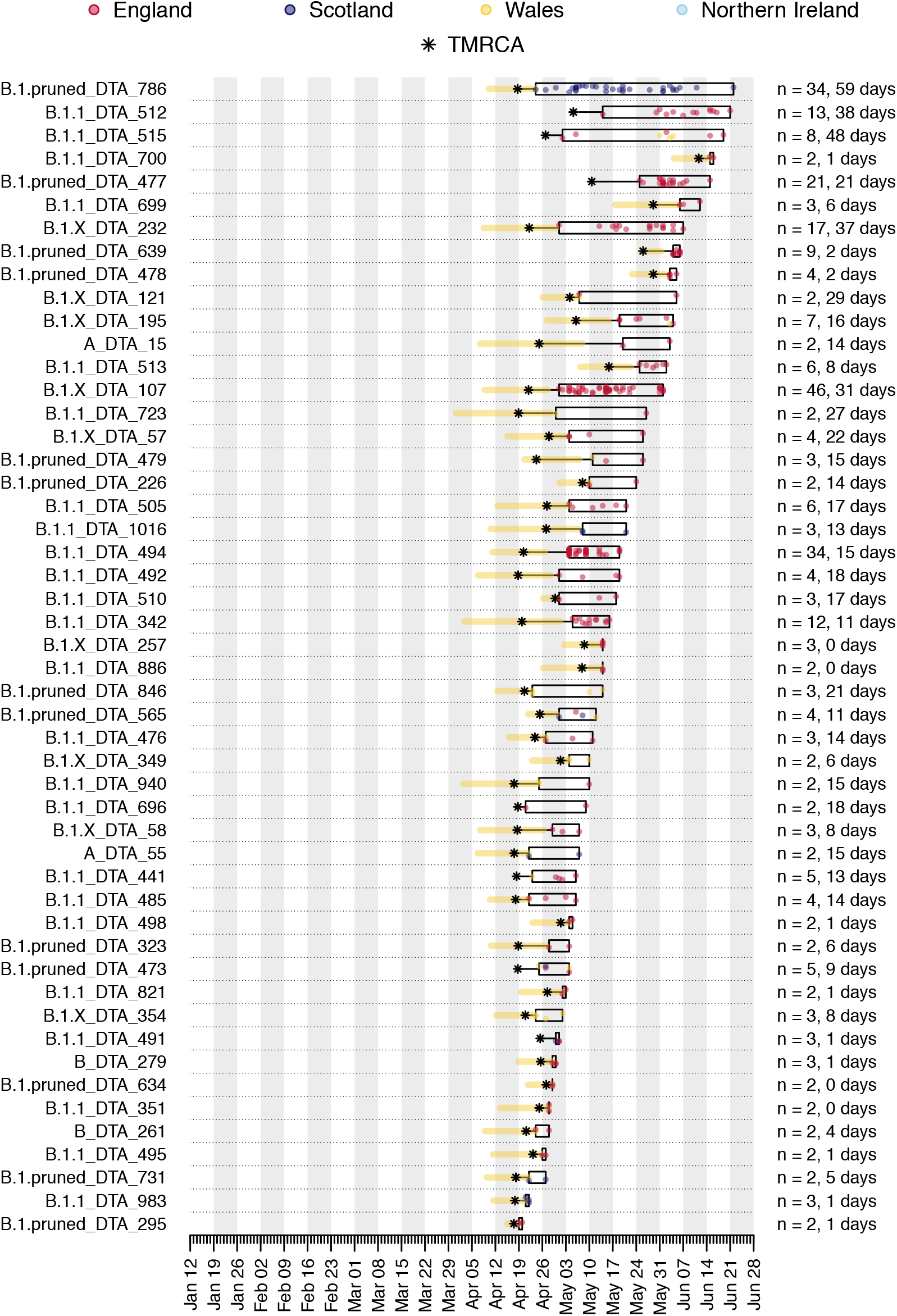
Illustration of the time course of the 50 most recent (by TMRCA) UK transmission lineages in our dataset. See Figure S4 caption for details.

**Fig. S7.**
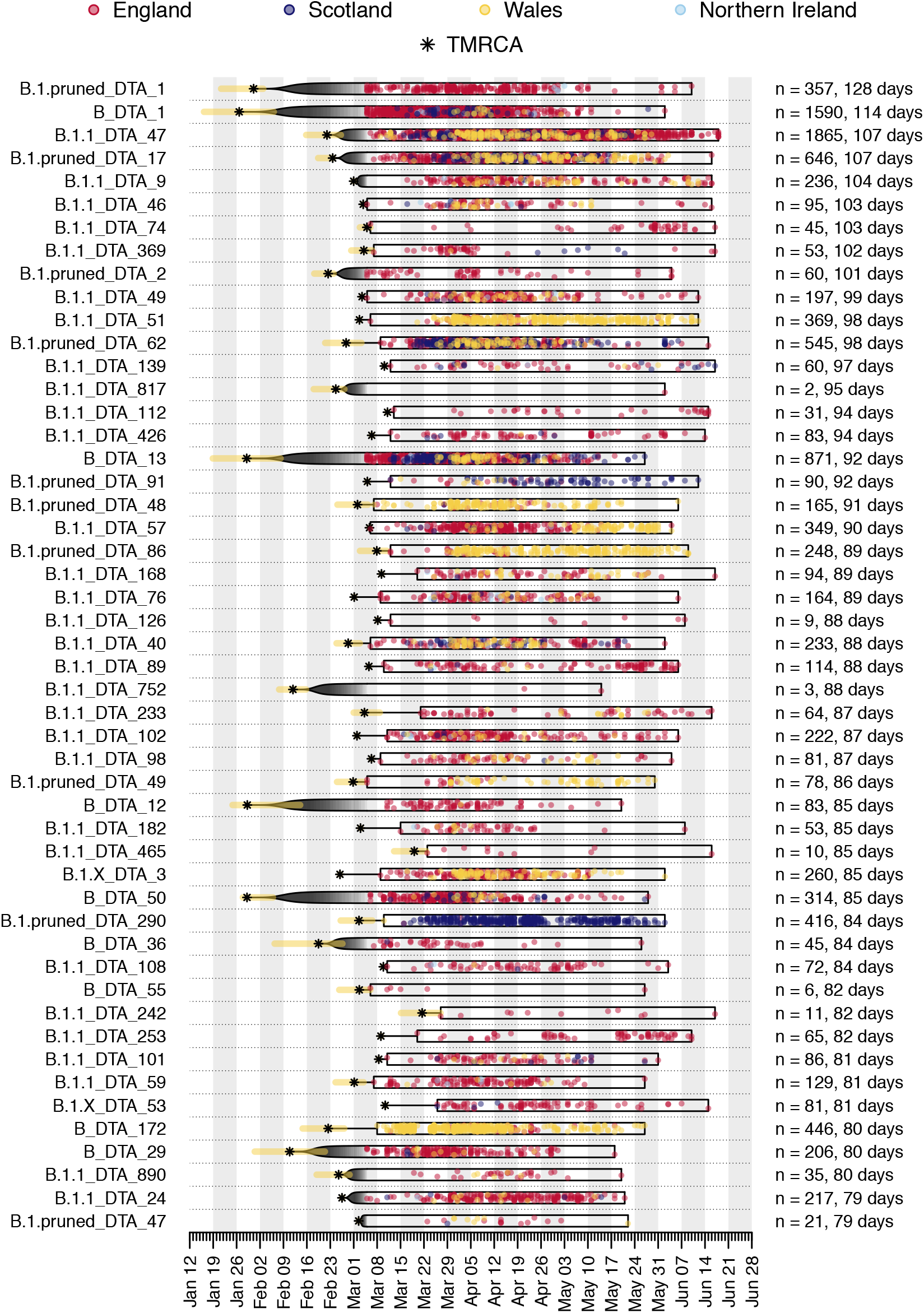
Illustration of the time course of the 50 UK transmission lineages with the longest sampling duration in our dataset. See Figure S4 caption for details.

**Fig. S8.**
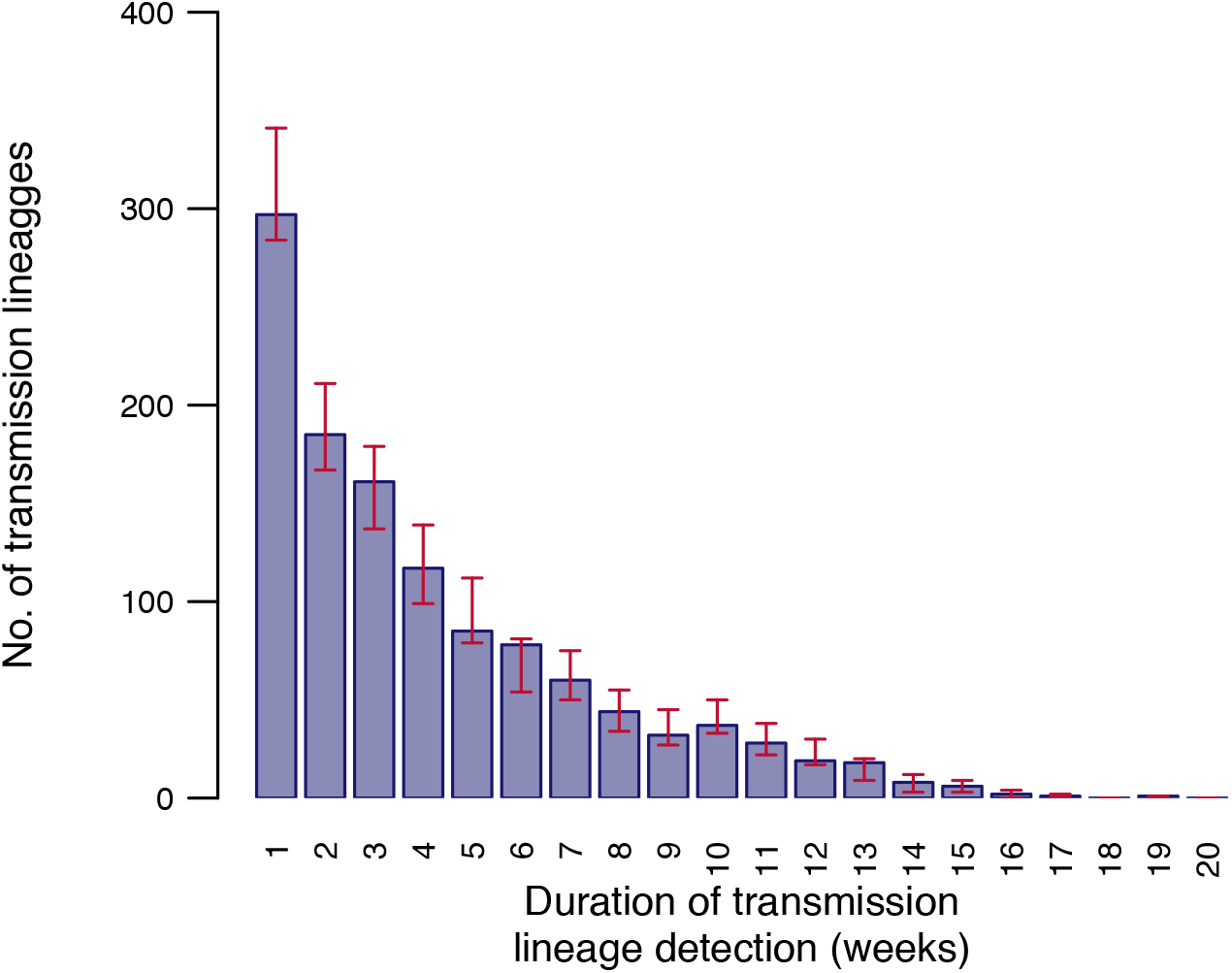
Distribution of UK transmission lineage sampling durations, aggregated by week. Blue bars show the number of transmission lineages that were observed over different durations in the MCC tree. Red bars show 95% HPD intervals for these numbers across the posterior tree distribution.

**Fig. S9.**
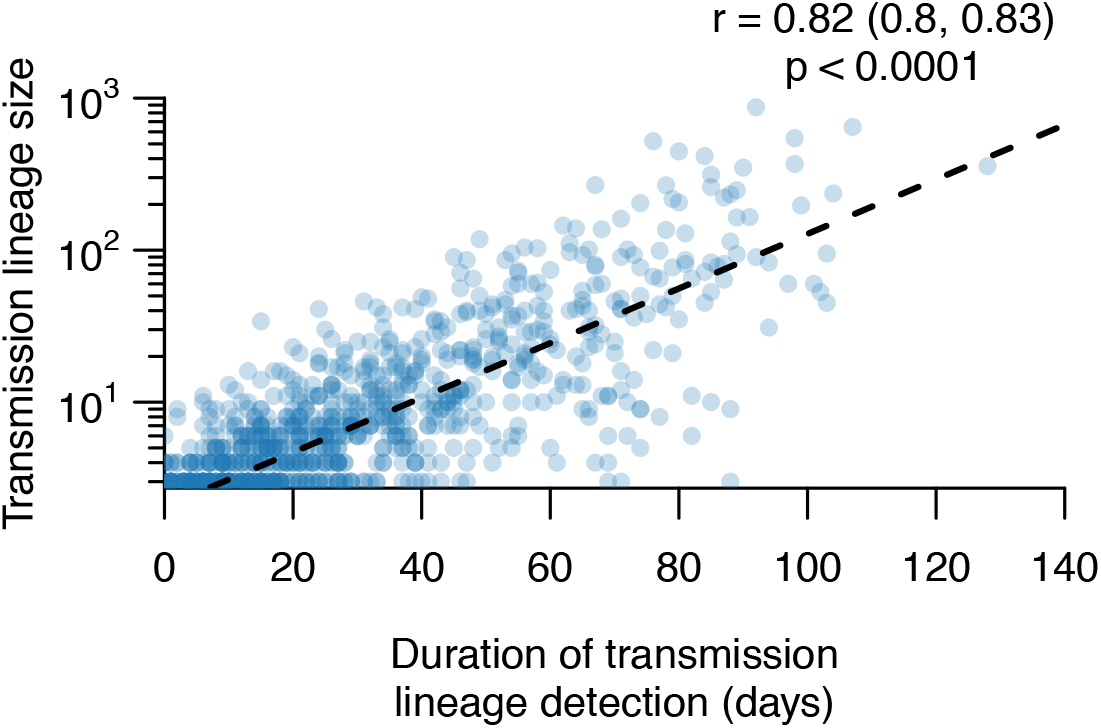
Scatterplot showing the strong relationship between UK transmission lineage size and sampling duration. The Pearson correlation coefficient, 95% CI and p-value are shown.

**Fig. S10.**
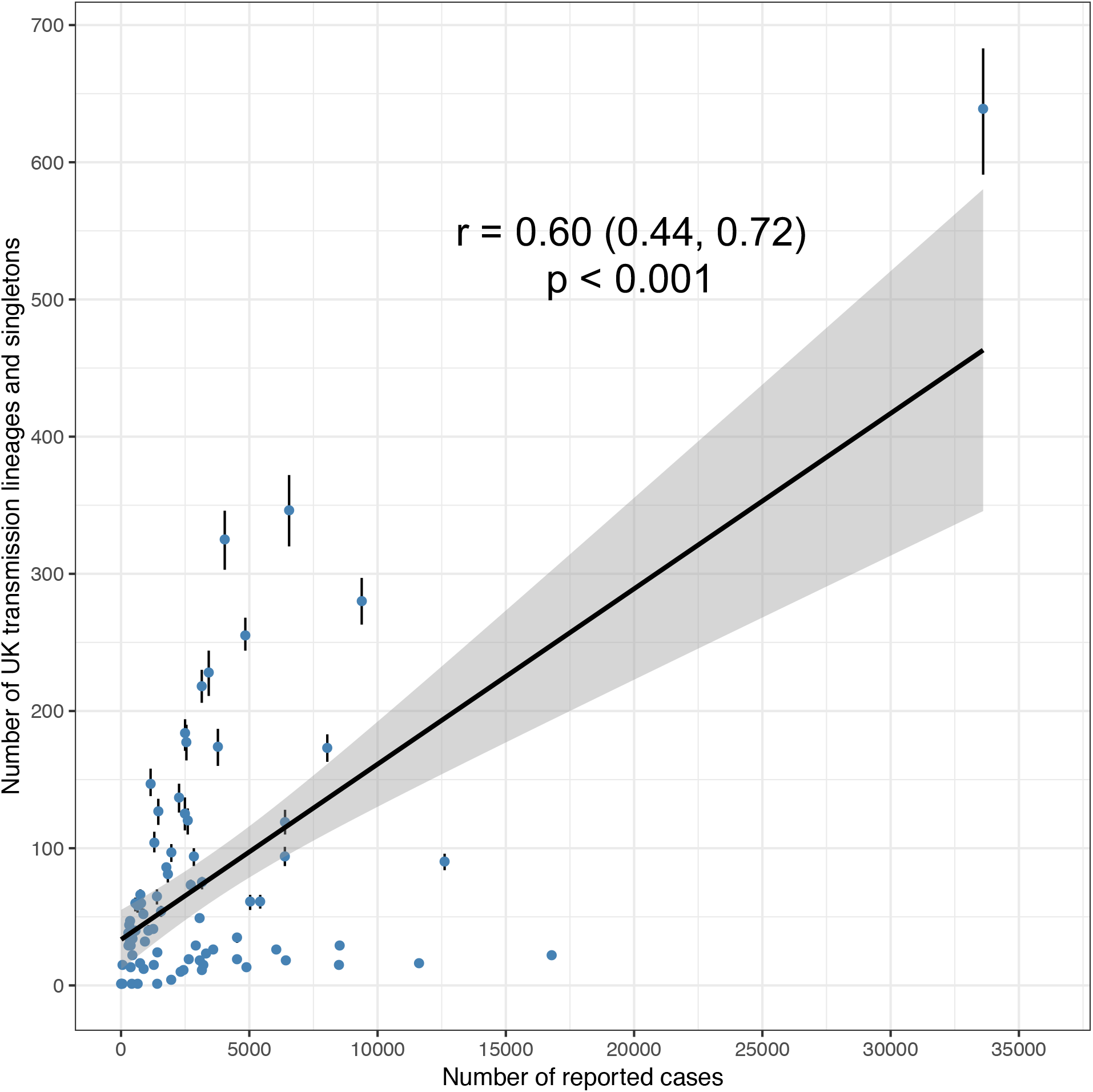
Scatterplot showing, for each geographic region, the relationship between the number of reported cases up to 26th June 2020 in that region and number of distinct UK transmission lineages and singletons detected in the region. Points show median estimates and error bars 95% HPDs from the posterior distribution of trees.

**Fig. S11.**
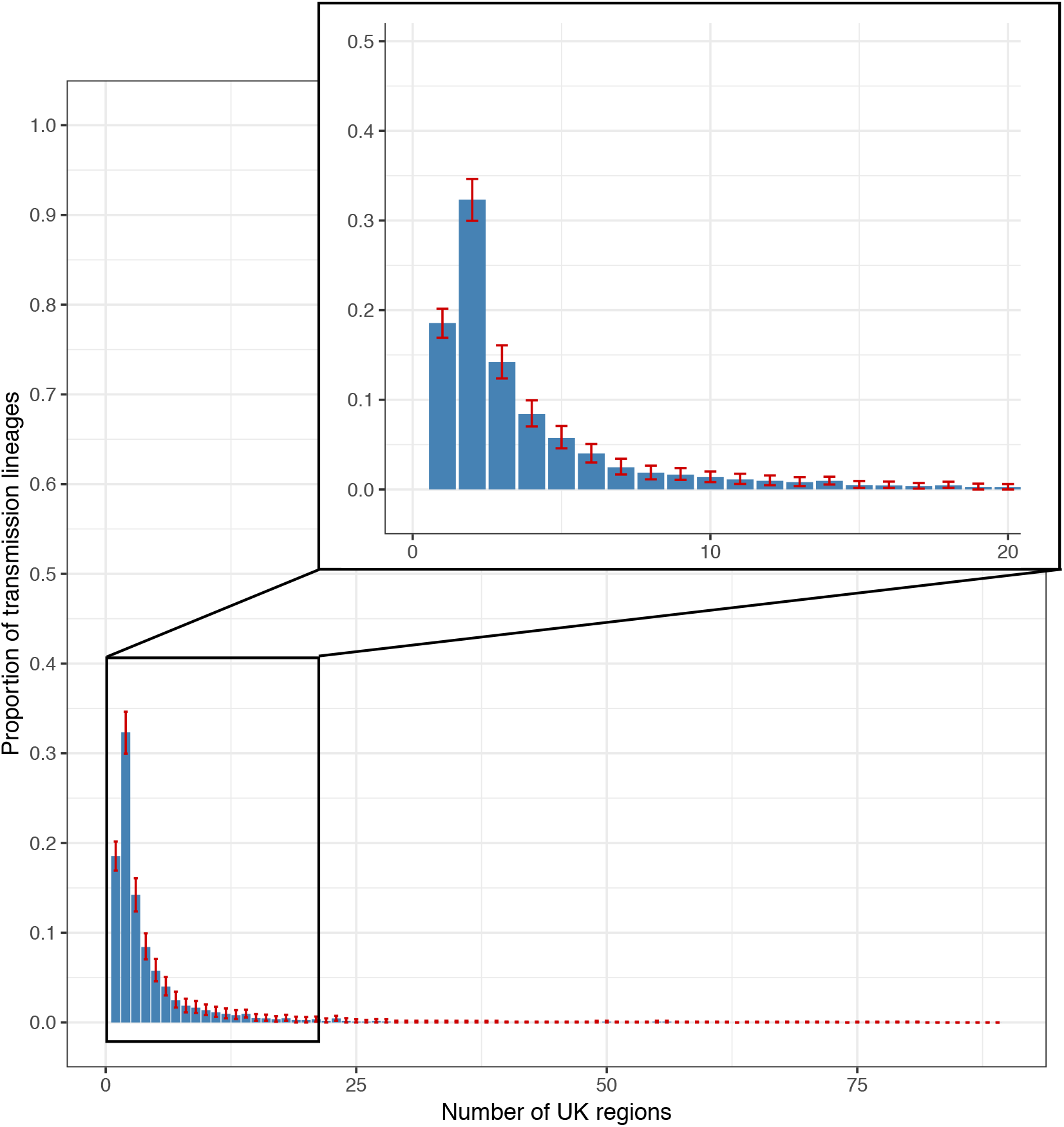
Geographic range size distribution of UK transmission lineages. Plot shows the distribution of the number of geographic regions in which each UK transmission lineage was sampled. Bars represent median proportions across the posterior distribution of trees and red bars show the 95% HPD intervals.

**Fig. S12.**
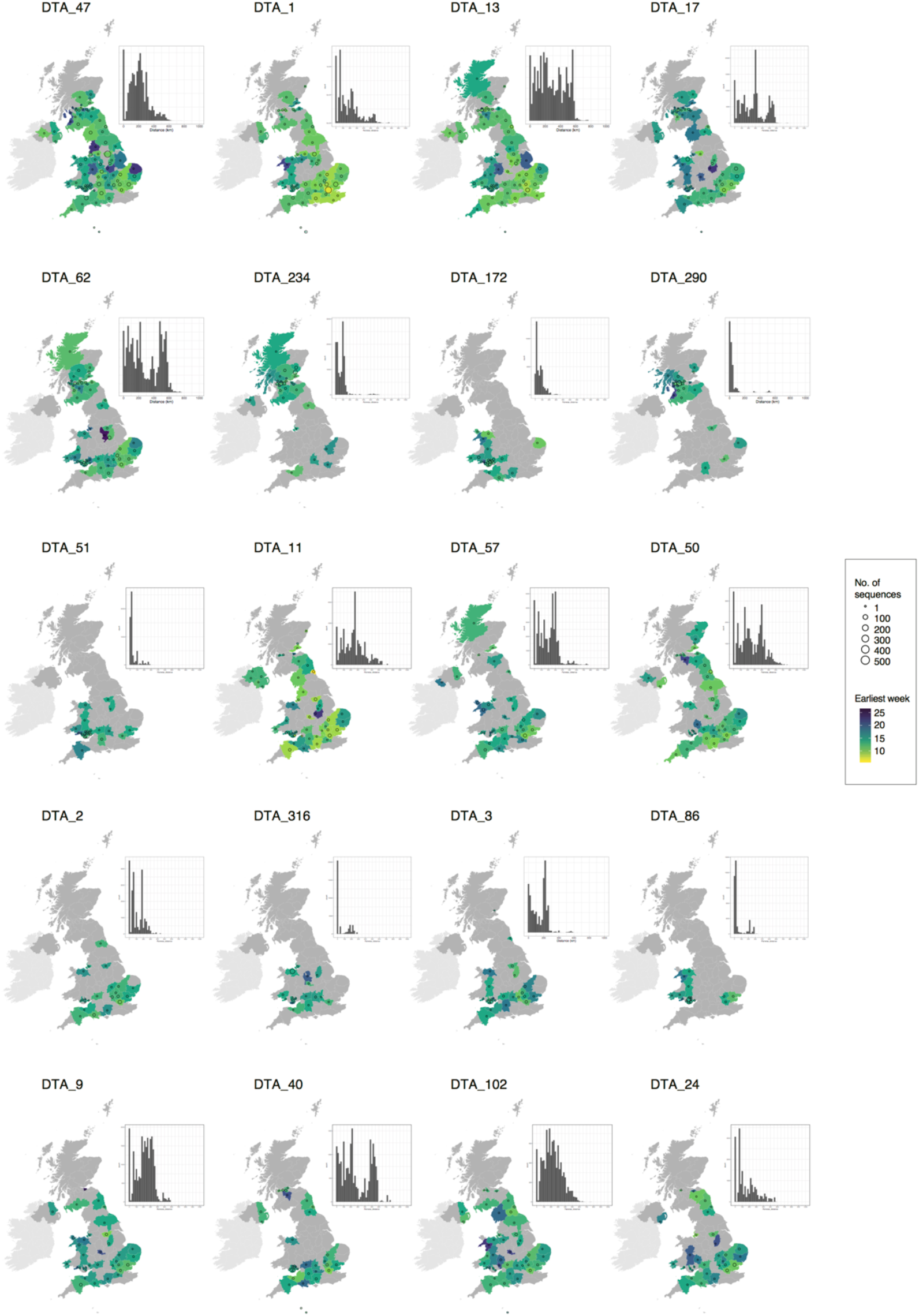
Spatial distribution of the twenty largest UK transmission lineages. Colours represent the week of the first detected genome in the transmission lineage in each location. Circles show the number of sampled genomes per location. Insets show the distribution of geographic distances for all sequence pairs within the lineage.

**Fig. S13.**
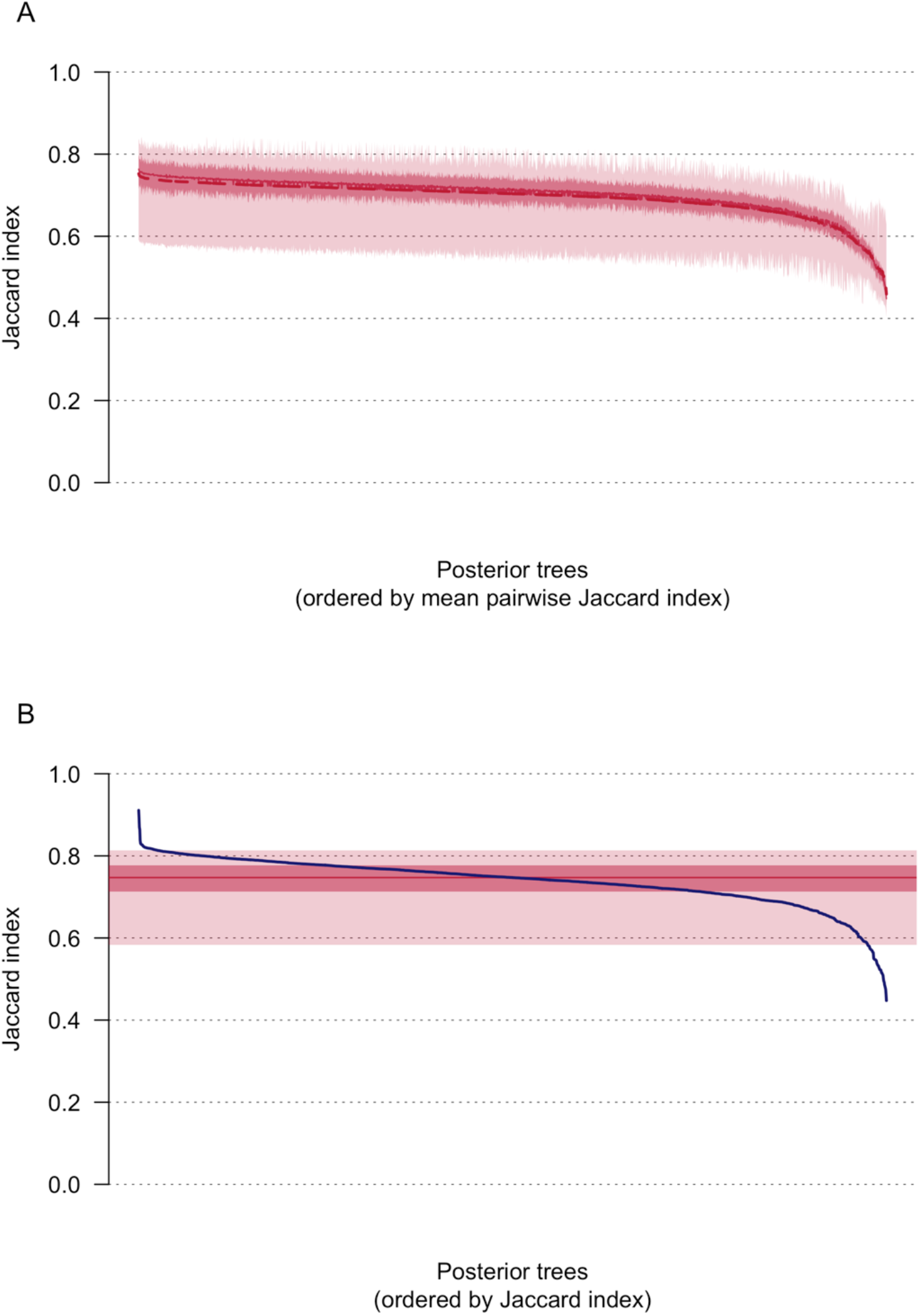
**(A)** Median (solid line) and mean (dashed line) Jaccard indices comparing the classification of UK genomes into transmission lineages and singletons on each of the 2000 posterior trees to the 1999 other trees. Dark shading shows the interquartile range and lighter shading the 95% CI. **(B)** Jaccard indices comparing the classification of UK genomes into transmission lineages and singletons on the MCC trees to each of the 2000 posterior trees (blue line). The solid red line indicates the median Jaccard index, dark shading the interquartile range and lighter shading the 95% CI.

**Fig. S14.**
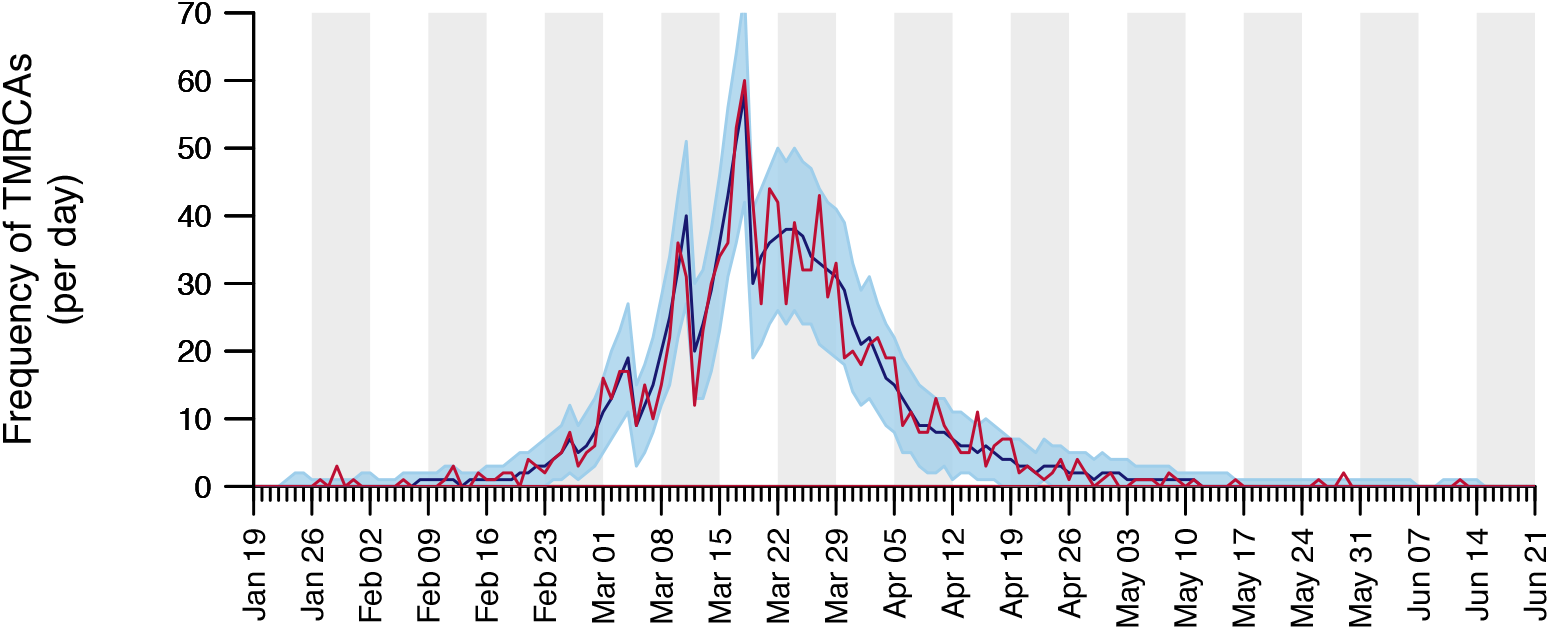
Comparison between the number of UK transmission lineage TMRCAs on each date in the MCC trees (red line) and across the 2000 posterior trees (median = blue line, 95% HPD interval = blue shading). Unevenness in this distribution is mostly likely caused by the phylogenetic constraints imposed by the sequence sampling times.

**Fig. S15.**
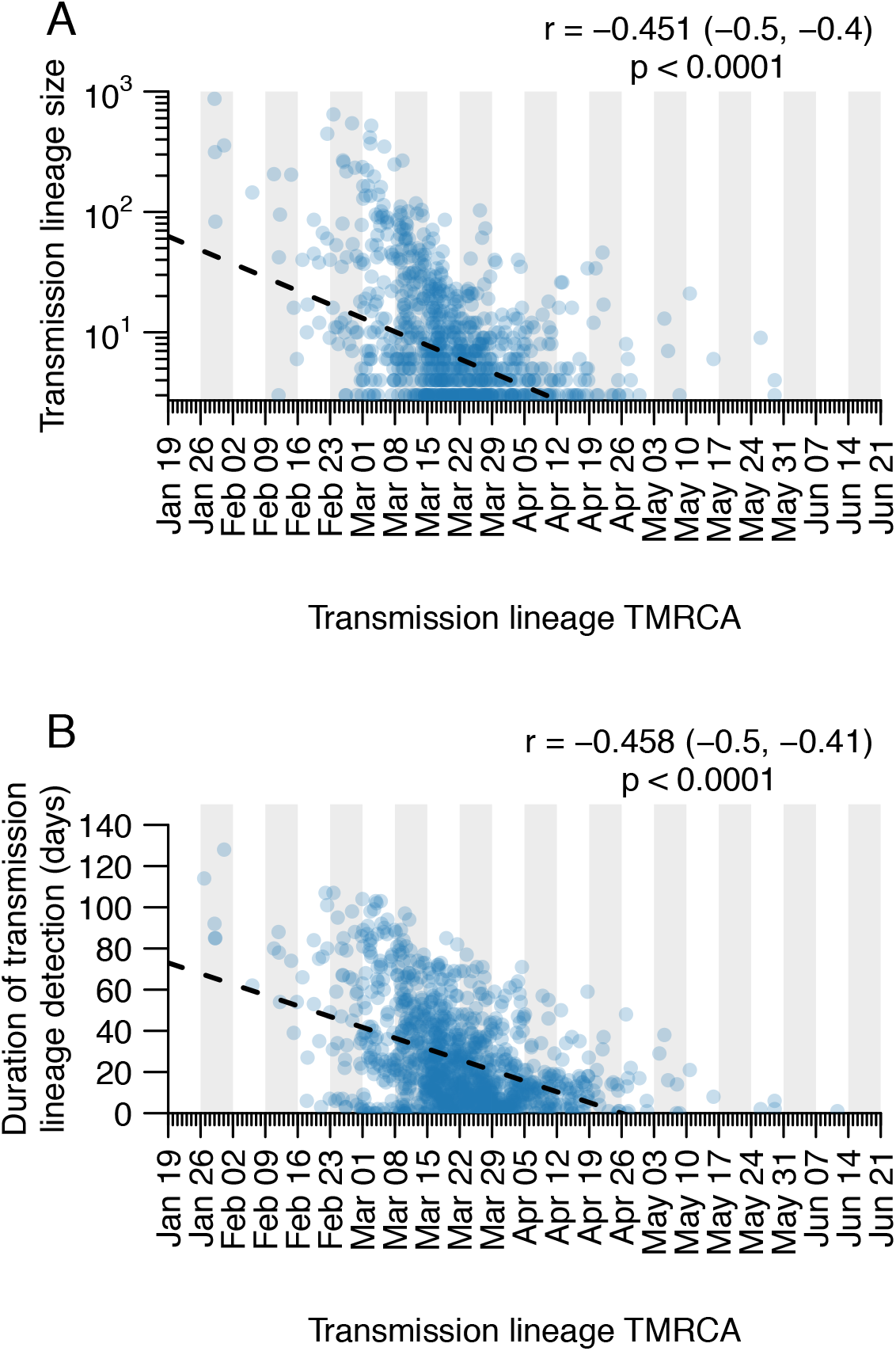
Scatterplots showing the relationship between (A) UK transmission lineage size and lineage TMRCA and between (B) UK transmission lineage sampling duration and lineage TMRCA. Pearson correlation coefficients, 95% CIs and p-values are shown in the top-right corners.

**Fig. S16.**
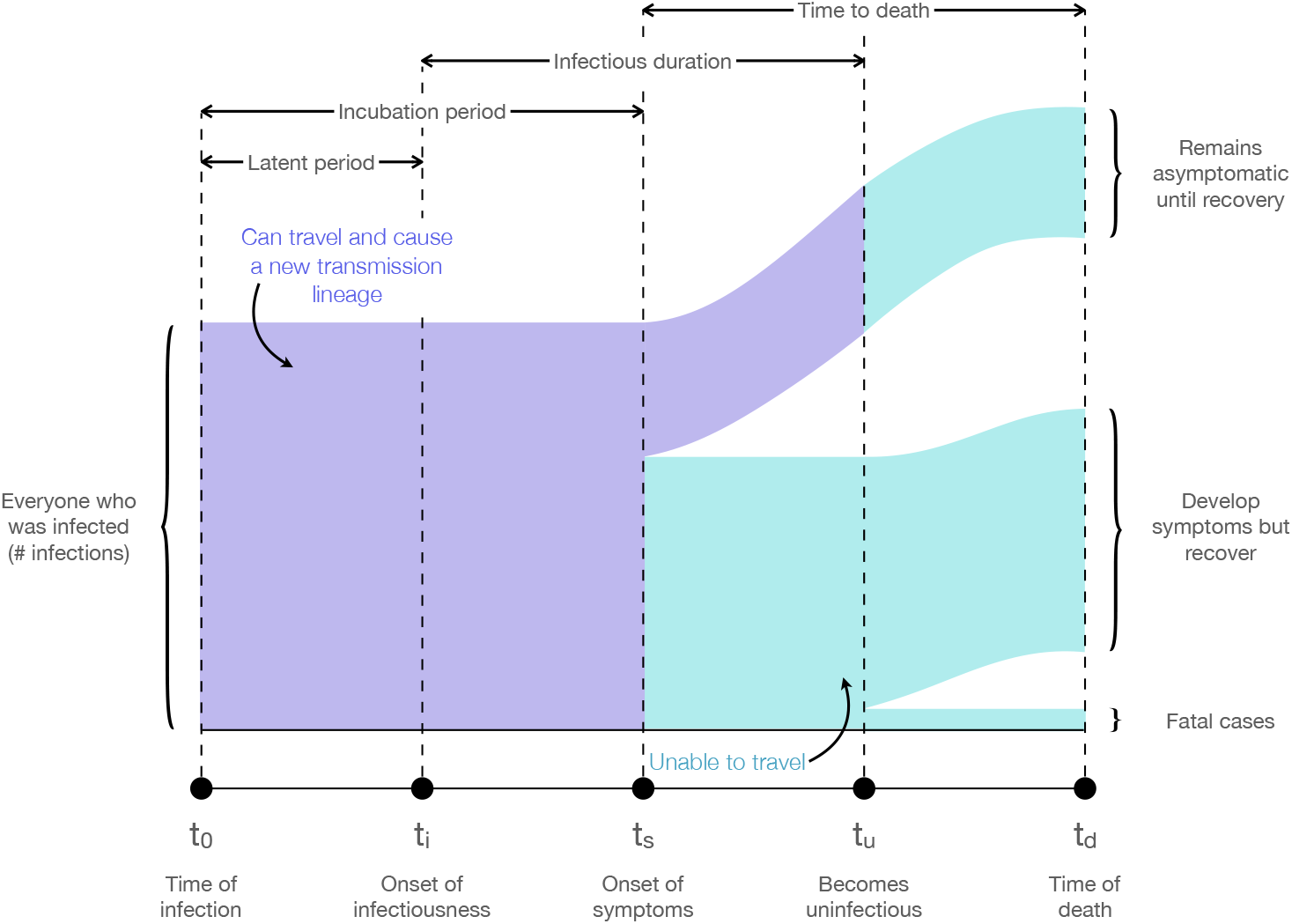
Sankey diagram showing the assumptions about the natural progression of a SARS-CoV-2 infection used in the estimation of global infectious cases. Infected individuals in the purple areas are potential initiators of a transmission lineage (PITL), but once they have progressed to the cyan areas they are assumed to no longer be capable of initiating a transmission lineage. We used the proportional flow through this diagram to estimate the total number of PITL through time given the number of COVID-19 associated deaths on each day.

**Fig. S17.**
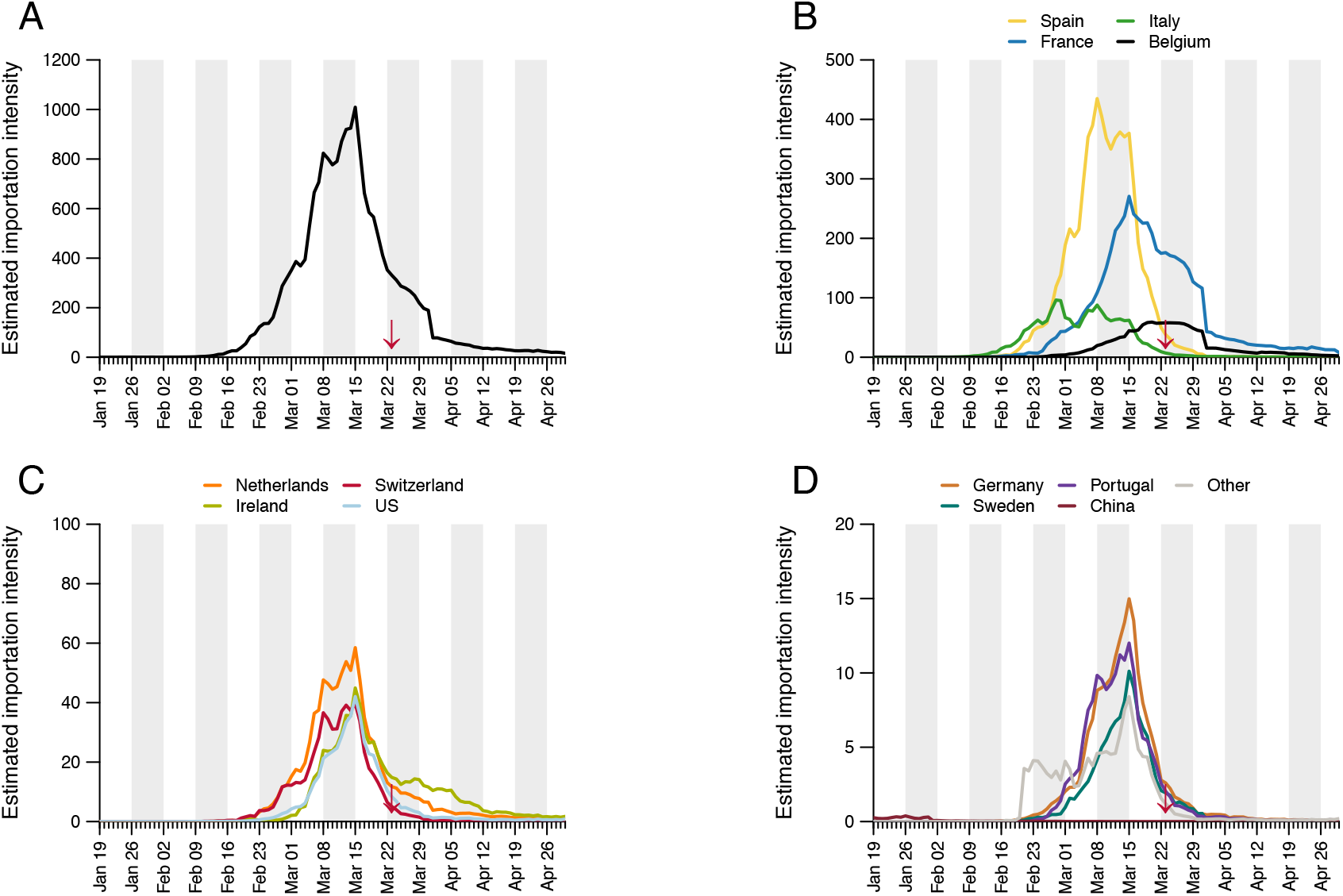
Estimated importation intensity (EII) curves for the 12 countries estimated to have contributed the most importations to the UK epidemic (see **Table S4**). Panel A shows the EII for all countries. The red arrows indicate the start of the UK lockdown.

**Fig S18.**
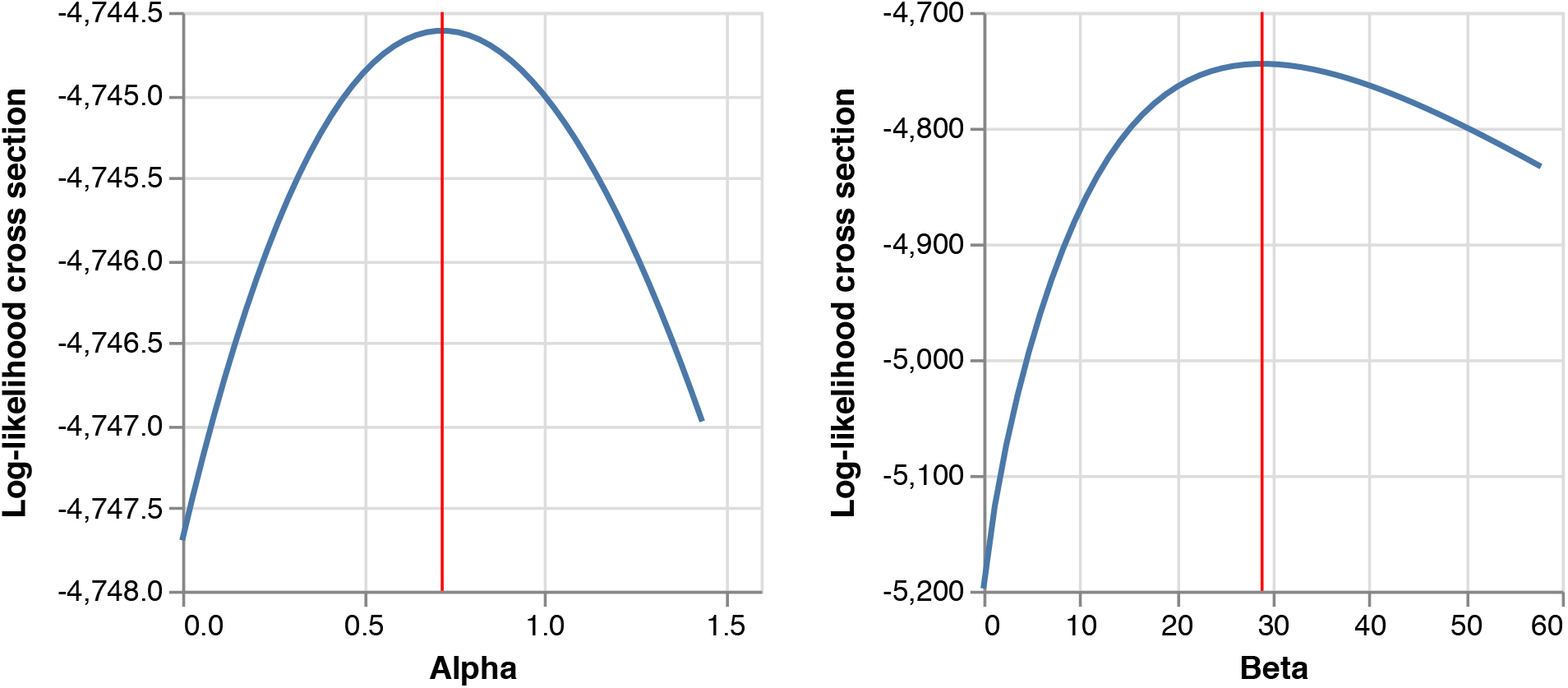
Log-likelihood function cross-section plots for possible parameter values of α and β inferred from genomic data, conditional on daily importation probabilities derived from the estimated importation intensities (EIIs). The maximum likelihood estimate (MLE) for each parameter is shown in red.

**Fig. S19.**
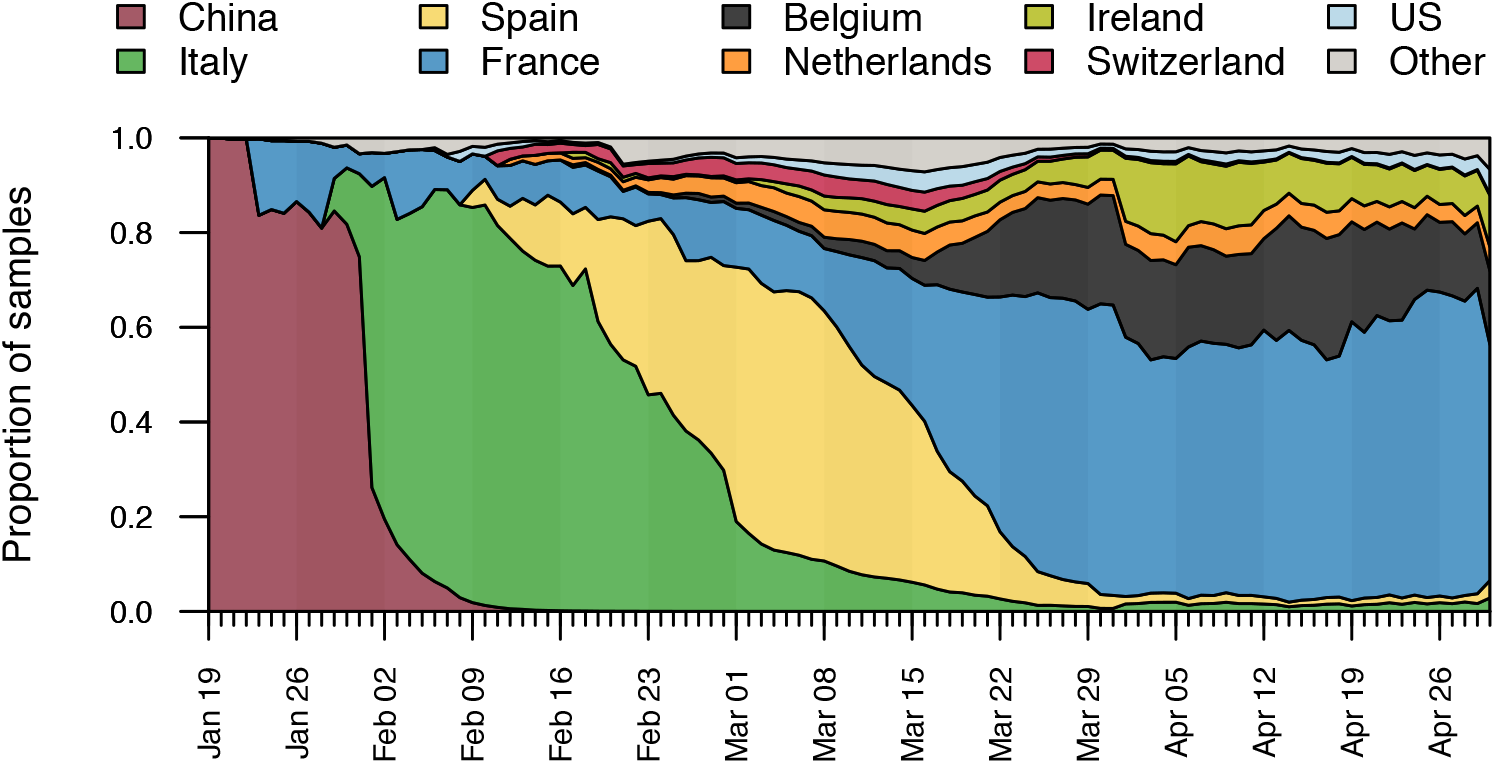
The estimated proportion of importation events that are attributable to inbound travellers from each of several source countries over time.

**Fig. S20.**
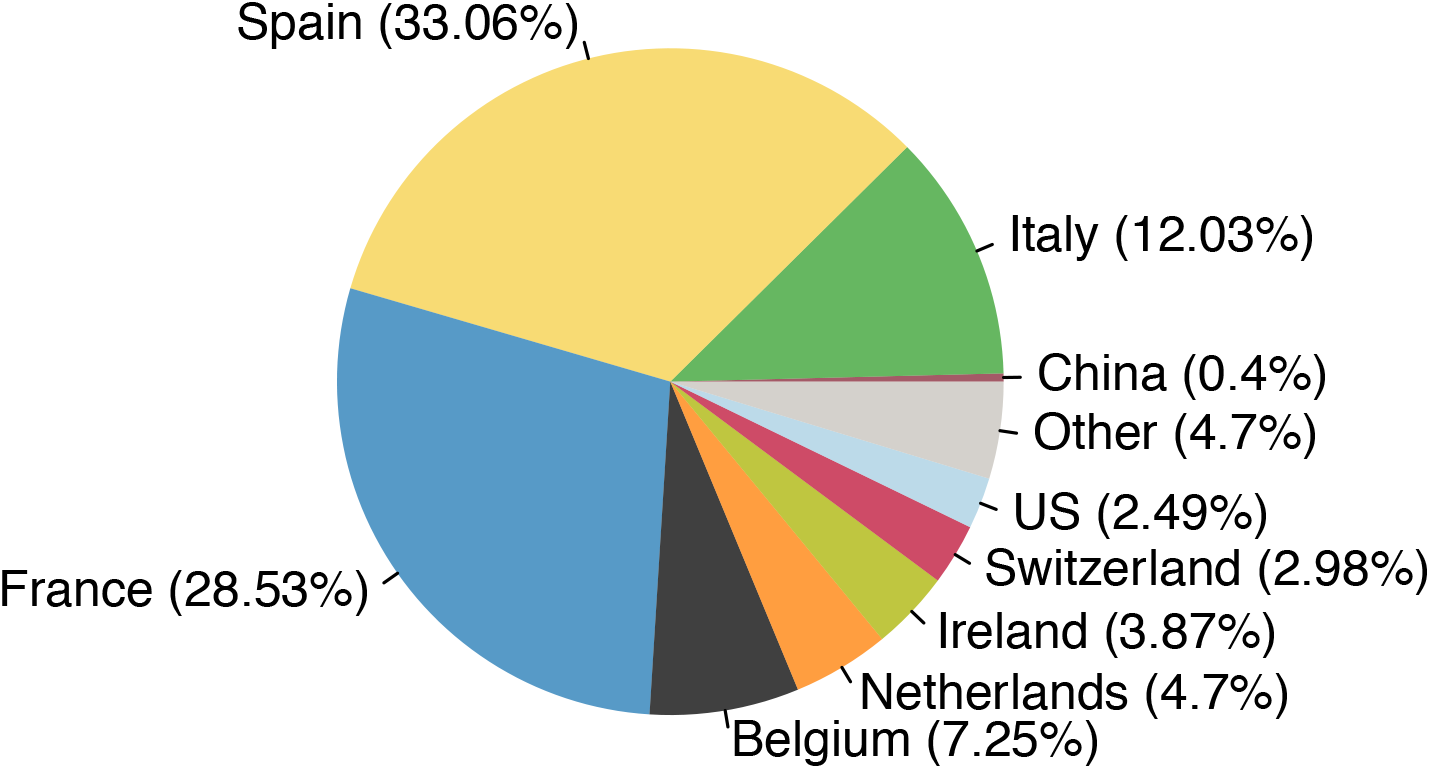
The estimated total fraction of importation events that are attributable to inbound travellers from each country.

**Table S1.** The number of location state transitions (non-UK to UK and vice-versa) taken across the set of 2000 posterior trees, as well as the total number of transmission lineages and singletons inferred across the set of 2000 posterior trees and the MCC trees. Numbers are given for the whole dataset and for each individual subtree.

| Lineages | Non-UK to UK state transitions (median, 95% HPD) | UK to non-UK state transitions (median, 95% HPD) | Transmission lineages and singletons (median, 95% HPD) | Transmission lineages and singletons in MCC tree |
| --- | --- | --- | --- | --- |
| Total | 2968<br>[2829-3103] | 1468<br>[1362-1566] | 2918<br>[2773-3048] | 2829 |
| A | 74<br>[67-80] | 3 [0-7] | 74 [67-80] | 69 |
| B | 372 [333-419] | 446 [402-485] | 365 [326-409] | 360 |
| B.1.1 | 1143 [1023-1258] | 666 [584-749] | 1115 [992-1230] | 1074 |
| B.1.pruned | 977 [925-1026] | 283 [245-321] | 964 [914-1015] | 943 |
| B.1.X | 398 [374-422] | 70 [49-93] | 396 [370-419] | 383 |

HPD – highest posterior density interval

**Table S2.** Estimated importation lags for UK transmission lineages of different sizes. Importation lag is the waiting time between importation date and the TMRCA of the sampled genomes in the transmission lineage (see **Fig. S2**). Detection lag is the waiting time from the importation date to the sampling time of the oldest (first) sampled genome in the transmission lineage (see **Fig. S2**).

| Lineages of size | No. of lineages | Importation lag (mean ± SD) | Importation lag (median, IQR) | Detection lag (mean ± SD) | Detection lag (median, IQR) |
| --- | --- | --- | --- | --- | --- |
| All | 1179 | 8.22 ± 5.21 | 7.95<br>[3.35-15.18] | 14.13 ± 5.61 | 14<br>[10-18] |
| 2 to 10 | 880 | 10.37 ± 4.24 | 10.36<br>[6.5-15.18] | 15.49 ± 5 | 16<br>[12-18] |
| 11 to 100 | 261 | 2.07 ± 0.74 | 2.03<br>[1.41-2.65] | 9.96 ± 4.92 | 9<br>[6-13] |
| 101 to 1000 | 36 | 0.87 ± 0.08 | 0.86<br>[0.81-0.93] | 11.08 ± 8.03 | 8.5<br>[5.75-15] |
| > 1000 | 2 | 0.74 ± 0 | 0.74<br>[0.74-0.74] | 12.5 ± 2.12 | 12.5<br>[11.75-13.25] |

SD – standard deviation
IQR – interquartile range

**Table S3.** Estimated importation and detection lags for UK transmission lineages ordered by importation date and aggregated by epi-week. Importation lag is the waiting time between importation date and the TMRCA of the sampled genomes in the transmission lineage (see **Fig. S2**). Detection lag is the waiting time from the importation date to the sampling time of the oldest (first) sampled genome in the transmission lineage (see **Fig. S2**). All statistics show means and standard deviations computed from the MCC trees.

| **Week starting** | **Epi-week** | **Estimated no. of importations** | **Lineage sizes (mean ± SD)** | **Importation lag (mean ± SD)** | **Detection lag (mean ± SD)** |
| --- | --- | --- | --- | --- | --- |
| **Jan-05** | 2 | 0 | - | - | - |
| **Jan-12** | 3 | 0 | - | - | - |
| **Jan-19** | 4 | 0 | - | - | - |
| **Jan-26** | 5 | 6 | 536.33 ± 598.96 | 2.42 ± 3.89 | 20.83 ± 11.81 |
| **Feb-02** | 6 | 2 | 73.5 ± 101.12 | 8.05 ± 10.08 | 20 ± 2.83 |
| **Feb-09** | 7 | 14 | 42.36 ± 73.45 | 8.72 ± 6.81 | 19.07 ± 4.41 |
| **Feb-16** | 8 | 45 | 63.4 ± 282.84 | 8.92 ± 5.8 | 14.27 ± 5.28 |
| **Feb-23** | 9 | 80 | 44.05 ± 108.48 | 7.82 ± 5.85 | 13.43 ± 6.46 |
| **Mar-01** | 10 | 206 | 26.53 ± 67.98 | 9.07 ± 5.64 | 14.34 ± 6.65 |
| **Mar-08** | 11 | 335 | 14.14 ± 25.04 | 7.87 ± 5.18 | 14.11 ± 5.39 |
| **Mar-15** | 12 | 235 | 8.32 ± 10.59 | 8.14 ± 4.91 | 13.54 ± 4.88 |
| **Mar-22** | 13 | 120 | 9.26 ± 13.88 | 7.78 ± 4.67 | 13.47 ± 4.78 |
| **Mar-29** | 14 | 71 | 5.96 ± 6.45 | 8.92 ± 4.62 | 14.77 ± 5.35 |
| **Apr-05** | 15 | 31 | 6.87 ± 6.59 | 7.92 ± 4.09 | 15.06 ± 5.83 |
| **Apr-12** | 16 | 15 | 6.6 ± 8.58 | 9.38 ± 4.67 | 15.73 ± 5.01 |
| **Apr-19** | 17 | 10 | 12.4 ± 15.49 | 7.9 ± 5.74 | 13.9 ± 3.84 |
| **Apr-26** | 18 | 3 | 7.67 ± 5.03 | 6.05 ± 3.85 | 15.33 ± 3.06 |
| **May-03** | 19 | 1 | 21 | 2.1 | 16 |
| **May-10** | 20 | 1 | 6 | 5.54 | 15 |
| **May-17** | 21 | 3 | 5.33 ± 3.21 | 7.41 ± 3.25 | 14.67 ± 2.89 |
| **May-24** | 22 | 1 | 2 | 15.18 | 19 |
| **May-31** | 23 | 0 | - | - | - |
| **Jun-07** | 24 | 0 | - | - | - |
| **Jun-14** | 25 | 0 | - | - | - |

**Table S4.** Number of observed importations in our dataset and the percentage of the total (1179) that can be attributed to the 40 countries inferred to be sources for the most importations.

| **Country** | **Observed importations** | **Percentage** |
| --- | --- | --- |
| Spain | 387.12 | 33.066 |
| France | 334.04 | 28.532 |
| Italy | 140.83 | 12.029 |
| Belgium | 84.88 | 7.25 |
| Netherlands | 55.08 | 4.705 |
| Ireland | 45.3 | 3.869 |
| Switzerland | 34.91 | 2.982 |
| US | 29.16 | 2.491 |
| Germany | 10.85 | 0.927 |
| Portugal | 9.56 | 0.817 |
| Sweden | 6.71 | 0.573 |
| China | 4.64 | 0.397 |
| Denmark | 3.84 | 0.328 |
| Austria | 3.48 | 0.297 |
| Romania | 2.24 | 0.191 |
| Norway | 1.95 | 0.167 |
| Poland | 1.28 | 0.109 |
| Canada | 1.08 | 0.093 |
| Turkey | 0.96 | 0.082 |
| Hungary | 0.95 | 0.081 |
| Czechia | 0.65 | 0.056 |
| Greece | 0.55 | 0.047 |
| United Arab Emirates | 0.3 | 0.026 |
| Israel | 0.27 | 0.023 |
| Finland | 0.25 | 0.022 |
| Iran | 0.22 | 0.019 |
| South Korea | 0.2 | 0.017 |
| Morocco | 0.18 | 0.015 |
| Brazil | 0.17 | 0.015 |
| Dominican Republic | 0.16 | 0.013 |
| Mexico | 0.08 | 0.007 |
| Serbia | 0.07 | 0.006 |
| Japan | 0.07 | 0.006 |
| Egypt | 0.06 | 0.005 |
| Malaysia | 0.05 | 0.004 |
| Pakistan | 0.05 | 0.004 |
| Moldova | 0.04 | 0.004 |
| Philippines | 0.04 | 0.003 |
| Russia | 0.03 | 0.003 |
| Ecuador | 0.03 | 0.003 |
| Other | 8.39 | 0.717 |

**Table S5.**
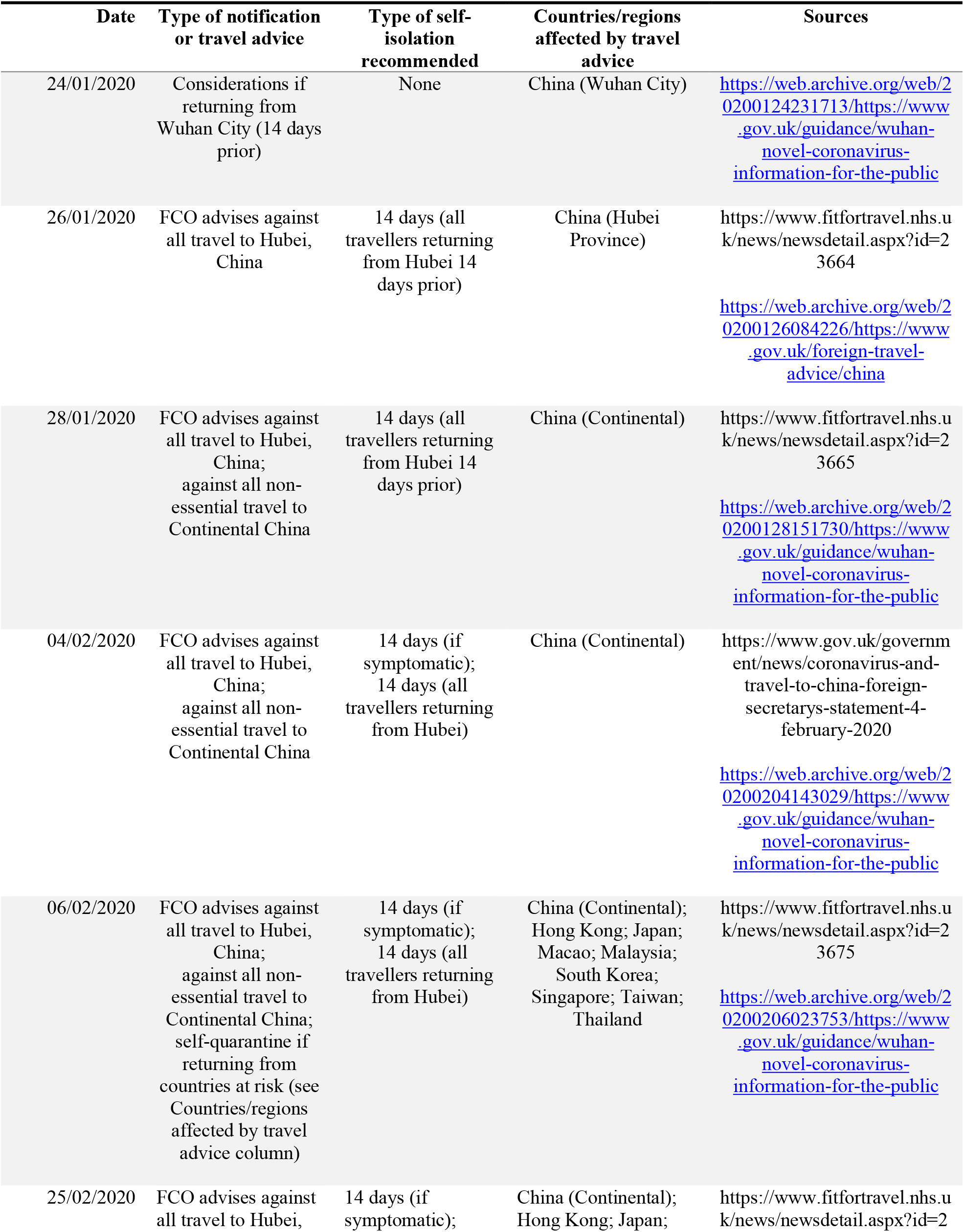
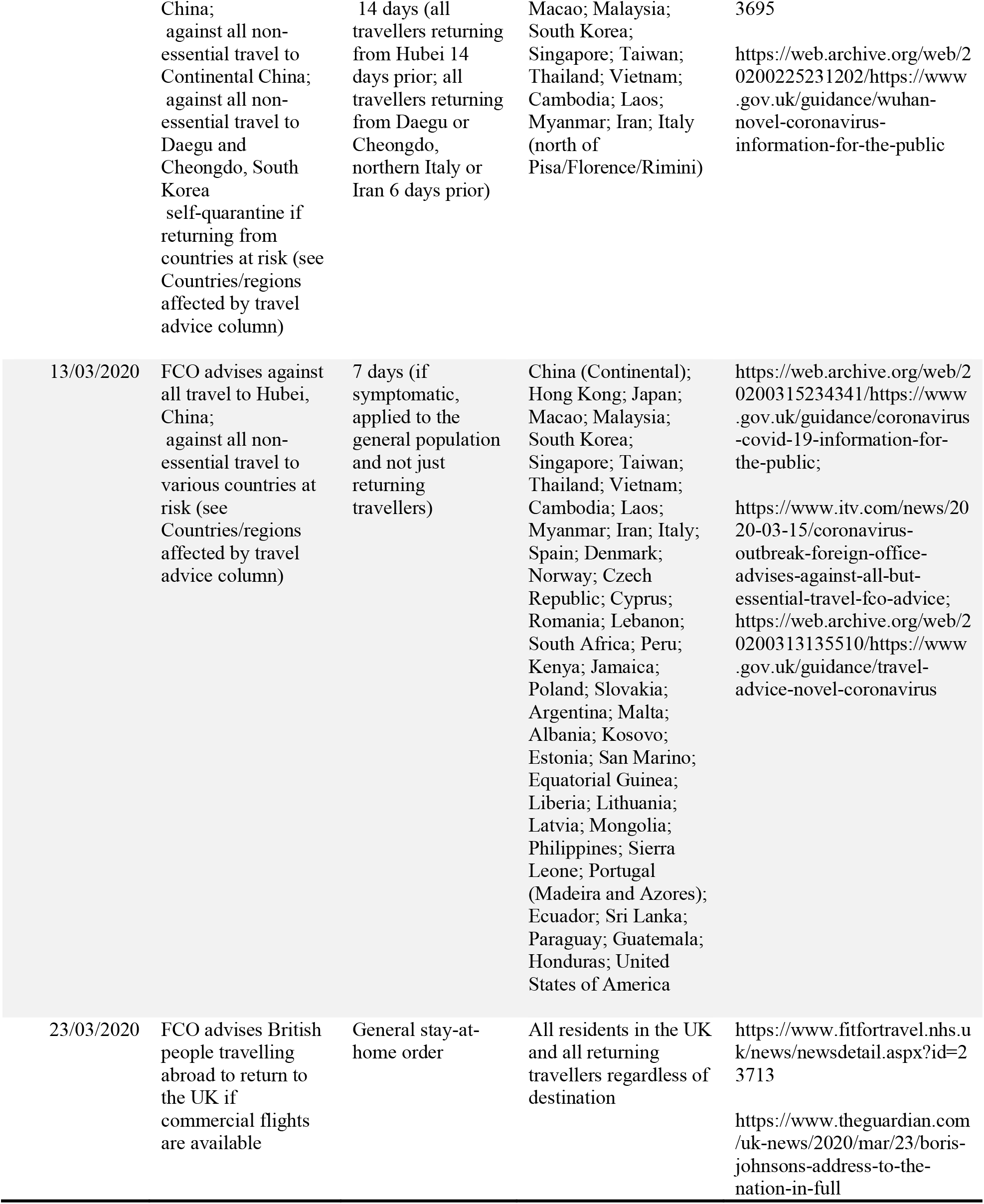
Summary of travel advice in the United Kingdom related to the COVID-19 pandemic from January to March.

## Notes

### Author Declarations

No such approval or exemption was necessary, as all data used are anonymised and publicly available.

