## Supplementary material for "Establishment & lineage dynamics of the SARS-CoV-2 epidemic in the UK": Full list of names and affiliations of COG-UK consortium members.

**Funding acquisition, leadership, supervision, metadata curation, project administration, samples, logistics, Sequencing, analysis, and Software and analysis tools:**
 Thomas R Connor ^33, 34^, and Nicholas J Loman ^15^.

**Leadership, supervision, sequencing, analysis, funding acquisition, metadata curation, project administration, samples, logistics, and visualisation:**

Samuel C Robson ^68^.

**Leadership, supervision, project administration, visualisation, samples, logistics, metadata curation and software and analysis tools:**

Tanya Golubchik ^27^.

**Leadership, supervision, metadata curation, project administration, samples, logistics sequencing and analysis:**

M. Estee Torok ^8, 10^.

**Project administration, metadata curation, samples, logistics, sequencing, analysis, and software and analysis tools:**

William L Hamilton ^8, 10^.

**Leadership, supervision, samples logistics, project administration, funding acquisition sequencing and analysis:**

David Bonsall ^27^.

**Leadership and supervision, sequencing, analysis, funding acquisition, visualisation and software and analysis tools:**

Ali R Awan ^74^.

**Leadership and supervision, funding acquisition, sequencing, analysis, metadata curation, samples and logistics:**

Sally Corden^33^ .

**Leadership supervision, sequencing analysis, samples, logistics, and metadata curation:** Ian Goodfellow ^11^.

**Leadership, supervision, sequencing, analysis, samples, logistics, and Project administration:**

Darren L Smith ^60, 61^.

**Project administration, metadata curation, samples, logistics, sequencing and analysis:**
Martin D Curran ^14^, and Surendra Parmar ^14^**.**

**Samples, logistics, metadata curation, project administration sequencing and analysis:**
James G Shepherd ^21^.

**Sequencing, analysis,	project administration, metadata curation and software and analysis tools:**
Matthew D Parker ^38^.

**Leadership, supervision, funding acquisition, samples, logistics, and metadata curation:**
Catherine Moore ^33^	.

**Leadership, supervision, metadata curation, samples, logistics, sequencing and analysis:**
Derek J Fairley^6, 88^, Matthew W Loose ^54^, and Joanne Watkins ^33^.

**Metadata curation, sequencing, analysis, leadership, supervision and software and analysis tools:**

Matthew Bull ^33^ , and Sam Nicholls ^15^	.

**Leadership, supervision, visualisation, sequencing, analysis and software and analysis tools:**

David M Aanensen ^1, 30^.

**Sequencing, analysis, samples, logistics, metadata curation, and visualisation:**
 Sharon Glaysher ^70^	.

**Metadata curation, sequencing, analysis, visualisation, software and analysis tools:**
 Matthew Bashton ^60^, and Nicole Pacchiarini ^33^.

**Sequencing, analysis, visualisation, metadata curation, and software and analysis tools**:
 Anthony P Underwood ^1, 30^.

**Funding acquisition, leadership, supervision and project administration:**
 Thushan I de Silva ^38^, and Dennis Wang ^38^**.**

**Project administration, samples, logistics, leadership and supervision**:

Monique Andersson^28^ , Anoop J Chauhan ^70^, Mariateresa de Cesare ^26^, Catherine Ludden ^1,3^ , and Tabitha W Mahungu ^91^.

**Sequencing, analysis, project administration and metadata curation:**
 Rebecca Dewar ^20^, and Martin P McHugh ^20^.

**Samples, logistics, metadata curation and project administration:**
Natasha G Jesudason ^21^, Kathy K Li MBBCh ^21^, Rajiv N Shah ^21^, and Yusri Taha ^66^.

**Leadership, supervision, funding acquisition and metadata curation:**
 Kate E Templeton ^20^**.**

**Leadership, supervision, funding acquisition, sequencing and analysis:**
Simon Cottrell ^33^, Justin O’Grady ^51^, Andrew Rambaut ^19^, and Colin P Smith^93^.

**Leadership, supervision, metadata curation	, sequencing and analysis:**
Matthew T.G. Holden ^87^, and Emma C Thomson ^21^.

**Leadership, supervision, samples, logistics and metadata curation**:
 Samuel Moses ^81, 82^.

**Sequencing, analysis, leadership, supervision, samples and logistics:**
Meera Chand ^7^, Chrystala Constantinidou ^71^, Alistair C Darby ^46^, Julian A Hiscox ^46^, Steve Paterson ^46^, and Meera Unnikrishnan ^71^**.**

**Sequencing, analysis, leadership and supervision and software and analysis tools:**
 Andrew J Page ^51^, and Erik M Volz ^96^.

**Samples, logistics, sequencing, analysis and metadata curation:**
Charlotte J Houldcroft ^8^, Aminu S Jahun ^11^, James P McKenna ^88^, Luke W Meredith ^11^, Andrew Nelson ^61^, Sarojini Pandey ^72^, and Gregory R Young ^60^.

**Sequencing, analysis, metadata curation, and software and analysis tools:**
 Anna Price ^34^, Sara Rey ^33^, Sunando Roy ^41^, Ben Temperton^49^, and Matthew Wyles ^38^.

**Sequencing, analysis, metadata curation and visualisation:**

Stefan	Rooke^19^, and Sharif Shaaban ^87^.

**Visualisation, sequencing, analysis and software and analysis tools:**
Helen Adams ^35^, Yann Bourgeois ^69^, Katie F Loveson ^68^, Áine O'Toole ^19^, and Richard Stark ^71^.

**Project administration, leadership and supervision:**

Ewan M Harrison ^1, 3^, David Heyburn ^33^, and Sharon J Peacock ^2, 3^

**Project administration and funding acquisition:**

David Buck ^26^ , and Michaela John^36^

**Sequencing, analysis and project administration:**

Dorota	Jamrozy ^1^, and Joshua Quick ^15^

**Samples, logistics, and project administration:**

Rahul Batra ^78^, Katherine L Bellis ^1, 3^, Beth Blane ^3^ , Sophia T Girgis ^3^, Angie Green ^26^, Anita Justice ^28^ , Mark Kristiansen ^41^ , and Rachel J Williams ^41^.

**Project administration, software and analysis tools:**

Radoslaw Poplawski^15^.

**Project administration and visualisation:**

Garry P Scarlett ^69^.

**Leadership, supervision, and funding acquisition:**

John A Todd ^26^, Christophe Fraser ^27^, Judith Breuer ^40,41^, Sergi Castellano ^41^, Stephen L Michell ^49^, Dimitris Gramatopoulos ^73^, and Jonathan Edgeworth ^78^.

**Leadership, supervision and metadata curation:**
Gemma L Kay ^51^.

**Leadership, supervision, sequencing and analysis:**
Ana da Silva Filipe ^21^ , Aaron R Jeffries ^49^, Sascha Ott ^71^, Oliver Pybus ^24^, David L Robertson ^21^, David A Simpson ^6^ , and Chris Williams ^33^.

**Samples, logistics, leadership and supervision:**

Cressida Auckland ^50^, John Boyes ^83^, Samir Dervisevic ^52^ , Sian Ellard ^49, 50^ , Sonia Goncalves^1^, Emma J Meader ^51^, Peter Muir ^2^, Husam Osman ^95^, Reenesh Prakash ^52^, Venkat Sivaprakasam ^18^, and Ian B Vipond ^2^.

**Leadership, supervision and visualisation**

Jane AH Masoli ^49, 50^.

**Sequencing, analysis and metadata curation**

Nabil-Fareed Alikhan ^51^, Matthew Carlile ^54^, Noel Craine ^33^, Sam T Haldenby ^46^, Nadine Holmes ^54^, Ronan A Lyons ^37^, Christopher Moore ^54^, Malorie Perry ^33^ , Ben Warne ^80^, and Thomas Williams ^19^.

**Samples, logistics and metadata curation:**

Lisa Berry ^72^, Andrew Bosworth ^95^ , Julianne Rose Brown ^40^, Sharon Campbell ^67^, Anna Casey ^17^, Gemma Clark ^56^, Jennifer Collins ^66^, Alison Cox ^43,^ ^44^ , Thomas Davis ^84^, Gary Eltringham ^66^, Cariad Evans ^38, 39^ , Clive Graham ^64^, Fenella Halstead ^18^, Kathryn Ann Harris ^40^, Christopher Holmes ^58^, Stephanie Hutchings ^2^ , Miren Iturriza-Gomara ^46^, Kate Johnson ^38, 39^, Katie Jones ^72^, Alexander J Keeley ^38^, Bridget A Knight ^49, 50^, Cherian Koshy^90^, Steven Liggett ^63^, Hannah Lowe ^81^ , Anita O Lucaci ^46^ , Jessica Lynch ^25, 29^ , Patrick C McClure ^55^, Nathan Moore ^31^ , Matilde Mori ^25, 29, 32^ , David G Partridge ^38, 39^ , Pinglawathee Madona ^43, 44^ , Hannah M Pymont ^2^, Paul Anthony Randell ^43, 44^ , Mohammad Raza ^38, 39^ , Felicity Ryan ^81^ , Robert Shaw ^28^, Tim J Sloan ^57^, and Emma Swindells ^65^ .

**Sequencing, analysis, Samples and logistics:**

Alexander Adams ^33^, Hibo Asad ^33^, Alec Birchley ^33^ , Tony Thomas Brooks ^41^, Giselda Bucca ^93^, Ethan Butcher ^70^, Sarah L Caddy ^13^, Laura G Caller ^2, 3, 12^ , Yasmin Chaudhry ^11^, Jason Coombes ^33^, Michelle Cronin ^33^, Patricia L Dyal ^41^, Johnathan M Evans ^33^, Laia Fina ^33^, Bree Gatica-Wilcox ^33^, Iliana Georgana ^11^, Lauren Gilbert ^33^ , Lee Graham ^33^, Danielle C Groves ^38^, Grant Hall ^11^, Ember Hilvers ^33^, Myra Hosmillo ^11^, Hannah Jones ^33^, Sophie Jones ^33^, Fahad A Khokhar ^13^ , Sara Kumziene-Summerhayes ^33^, George MacIntyre-Cockett ^26^, Rocio T Martinez Nunez ^94^, Caoimhe McKerr ^33^, Claire McMurray ^15^, Richard Myers ^7^, Yasmin Nicole Panchbhaya ^41^, Malte L Pinckert ^11^ , Amy Plimmer ^33^ , Joanne Stockton ^15^ , Sarah Taylor ^33^ , Alicia Thornton ^7^ , Amy Trebes ^26^ , Alexander J Trotter ^51^ ,Helena Jane Tutill ^41^ ,Charlotte A Williams ^41^ , Anna Yakovleva ^11^ and Wen C Yew ^62^.

**Sequencing, analysis and software and analysis tools:**
Mohammad T Alam ^71^, Laura Baxter ^71^, Olivia Boyd ^96^ , Fabricia F. Nascimento ^96^, Timothy M Freeman ^38^, Lily Geidelberg ^96^, Joseph Hughes ^21^, David Jorgensen ^96^, Benjamin B Lindsey ^38^, Richard J Orton ^21^ , Manon Ragonnet-Cronin ^96^ Joel Southgate ^33, 34,^ and Sreenu Vattipally ^21^.

**Samples, logistics and software and analysis tools:**

Igor Starinskij ^23^.

**Visualisation and software and analysis tools:**
Joshua B Singer ^21^ , Khalil Abudahab ^1, 30^, Leonardo de Oliveira Martins ^51^ , Thanh Le-Viet ^51^ ,Mirko Menegazzo ^30^ ,Ben EW Taylor ^1, 30^, and Corin A Yeats ^30^.

**Project Administration:**

Sophie Palmer	^3^, Carol M Churcher ^3^ , Alisha Davies ^33^, Elen De Lacy ^33^, Fatima Downing ^33^, Sue Edwards ^33^ , Nikki Smith ^38^ , Francesc Coll ^97^ , Nazreen F Hadjirin ^3^ and Frances Bolt ^44, 45^.

**Leadership and supervision:**

Alex Alderton^1^, Matt Berriman^1^, Ian G Charles ^51^, Nicholas Cortes ^31^, Tanya Curran ^88^, John Danesh^1^, Sahar Eldirdiri ^84^, Ngozi Elumogo ^52^, Andrew Hattersley ^49, 50^, Alison Holmes ^44, 45^, Robin Howe ^33^, Rachel Jones ^33^, Anita Kenyon ^84^, Robert A Kingsley ^51^, Dominic Kwiatkowski ^1, 9^, Cordelia Langford^1^, Jenifer Mason^48^, Alison E Mather ^51^, Lizzie Meadows ^51^, Sian Morgan ^36^, James Price ^44, 45^, Trevor I Robinson ^48^, Giri Shankar ^33^ , John Wain ^51^, and Mark A Webber ^51^.

**Metadata curation:**

Declan T Bradley ^5, 6^, Michael R Chapman ^1, 3, 4^ , Derrick Crooke ^28^ , David Eyre ^28^, Martyn Guest ^34^ , Huw Gulliver ^34^, Sarah Hoosdally ^28^, Christine Kitchen ^34^, Ian Merrick ^34^, Siddharth Mookerjee ^44, 45^, Robert Munn ^34^ , Timothy Peto ^28^, Will Potter ^52^, Dheeraj K Sethi ^52^, Wendy Smith ^56^ , Luke B Snell ^75, 94^, Rachael Stanley ^52^ , Claire Stuart ^52^ and Elizabeth Wastenge^20^.

**Sequencing and analysis:**

Erwan Acheson ^6^ , Safiah Afifi ^36^ , Elias Allara ^2, 3^ , Roberto Amato ^1^, Adrienn Angyal ^38^, Elihu Aranday-Cortes ^21^ , Cristina Ariani ^1^, Jordan Ashworth ^19^, Stephen Attwood ^24^, Alp Aydin ^51^, David J Baker ^51^, Carlos E Balcazar ^19^, Angela Beckett	^68^ Robert Beer ^36^, Gilberto Betancor ^76^, Emma Betteridge ^1^ , David Bibby ^7^ , Daniel Bradshaw^7^ , Catherine Bresner ^34^, Hannah E Bridgewater ^71^ , Alice Broos ^21^, Rebecca Brown ^38^ , Paul E Brown ^71^, Kirstyn Brunker ^22^ , Stephen N Carmichael ^21^ , Jeffrey K. J. Cheng ^71^, Dr Rachel Colquhoun ^19^, Gavin Dabrera ^7^ , Johnny Debebe ^54^, Eleanor Drury ^1^, Louis du Plessis ^24^ , Richard Eccles ^46^, Nicholas Ellaby ^7^, Audrey Farbos ^49^, Ben Farr ^1^, Jacqueline Findlay ^41^ , Chloe L Fisher ^74^, Leysa Marie Forrest ^41^, Sarah Francois ^24^, Lucy R. Frost ^71^, William Fuller^34^ , Eileen Gallagher ^7^, Michael D Gallagher ^19^ , Matthew Gemmell ^46^, Rachel AJ Gilroy ^51^, Scott Goodwin ^1^, Luke R Green ^38^, Richard Gregory ^46^ , Natalie Groves ^7^, James W Harrison ^49^, Hassan Hartman ^7^ , Andrew R Hesketh ^93^,Verity Hill ^19^, Jonathan Hubb ^7^, Margaret Hughes^46^ , David K Jackson ^1^ , Ben Jackson ^19^, Keith James ^1^ ,Natasha Johnson ^21^ ,Ian Johnston ^1^, Jon-Paul Keatley ^1^, Moritz Kraemer ^24^, Angie Lackenby ^7^, Mara Lawniczak ^1^ , David Lee ^7^, Rich Livett ^1^, Stephanie Lo ^1^, Daniel Mair ^21^, Joshua Maksimovic ^36^, Nikos Manesis ^7^ , Robin Manley ^49^, Carmen Manso ^7^, Angela Marchbank ^34^ , Inigo Martincorena ^1^ , Tamyo Mbisa ^7^, Kathryn McCluggage ^36^, JT McCrone ^19^, Shahjahan Miah ^7^ , Michelle L Michelsen ^49^, Mari Morgan ^33^, Gaia Nebbia ^78^,Charlotte Nelson ^46^ ,Jenna Nichols ^21^ ,Paola Niola ^41^ , Kyriaki Nomikou ^21^ ,Steve Palmer ^1^ , Naomi Park ^1^, Yasmin A Parr ^1^ , Paul J Parsons ^38^ , Vineet Patel ^7^ , Minal Patel ^1^ ,Clare Pearson ^2, 1^, Steven Platt ^7^ ,Christoph Puethe ^1^, Mike Quail ^1^,Jayna Raghwani ^24^ , Lucille Rainbow ^46^ ,Shavanthi Rajatileka ^1^, Mary Ramsay ^7^ , Paola C Resende Silva ^41, 42^, Steven Rudder 51, Chris Ruis ^3^ , Christine M Sambles ^49^, Fei Sang ^54^, Ulf Schaefer^7^, Emily Scher ^19^, Carol Scott ^1^ ,Lesley Shirley ^1^, Adrian W Signell ^76^, John Sillitoe ^1^ ,Christen Smith ^1^ ,Dr Katherine L Smollett ^21^ ,Karla Spellman ^36^ ,Thomas D Stanton ^19^, David J Studholme ^49^ ,Grace Taylor-Joyce ^71^ ,Ana P Tedim ^51^, Thomas Thompson ^6^, Nicholas M Thomson ^51^, Scott Thurston^1^ , Lily Tong ^21^, Gerry Tonkin-Hill ^1^, Rachel M Tucker ^38^ , Edith E Vamos ^4^, Tetyana Vasylyeva^24^, Joanna Warwick-Dugdale ^49^ , Danni Weldon ^1^, Mark Whitehead ^46^, David Williams ^7^, Kathleen A Williamson ^19^,Harry D Wilson ^76^,Trudy Workman ^34^, Muhammad Yasir^51^, Xiaoyu Yu ^19^, and Alex Zarebski ^24^.

**Samples and logistics:**

Evelien M Adriaenssens ^51^, Shazaad S Y Ahmad ^2, 47^ , Adela Alcolea-Medina ^59, 77^, John Allan ^60^, Patawee Asamaphan ^21^, Laura Atkinson ^40^, Paul Baker ^63^, Jonathan Ball ^55^, Edward Barton^64^, Mathew A Beale^1^, Charlotte Beaver^1^, Andrew Beggs ^16^, Andrew Bell ^51^, Duncan J Berger ^1^, Louise Berry. ^56^, Claire M Bewshea ^49^, Kelly Bicknell ^70^, Paul Bird ^58^, Chloe Bishop ^7^ , Tim Boswell ^56^, Cassie Breen ^48^, Sarah K Buddenborg^1^, Shirelle Burton-Fanning ^66^ , Vicki Chalker ^7^, Joseph G Chappell ^55^, Themoula Charalampous ^78, 94^, Claire Cormie^3^, Nick Cortes^29, 25^, Lindsay J Coupland ^52^, Angela Cowell ^48^ , Rose K Davidson ^53^ , Joana Dias ^3^, Maria Diaz ^51^ , Thomas Dibling^1^, Matthew J Dorman^1^, Nichola Duckworth^57^, Scott Elliott^70^, Sarah Essex^63^, Karlie Fallon ^58^ , Theresa Feltwell ^8^, Vicki M Fleming ^56^, Sally Forrest ^3^, Luke Foulser^1^, Maria V Garcia-Casado^1^, Artemis Gavriil ^41^, Ryan P George ^47^, Laura Gifford ^33^, Harmeet K Gill ^3^, Jane Greenaway ^65^, Luke Griffith^53^, Ana Victoria Gutierrez^51^, Antony D Hale ^85^, Tanzina Haque ^91^, Katherine L Harper ^85^, Ian Harrison ^7^ , Judith Heaney ^89^, Thomas Helmer ^58^, Ellen E Higginson^3^ , Richard Hopes ^2^, Hannah C Howson-Wells ^56^, Adam D Hunter ^1^, Robert Impey ^70^, Dianne Irish-Tavares ^91^, David A Jackson^1^ , Kathryn A Jackson ^46^, Amelia Joseph ^56^, Leanne Kane ^1^, Sally Kay ^1^, Leanne M Kermack ^3^, Manjinder Khakh ^56^, Stephen P Kidd ^29, 25,31^, Anastasia Kolyva ^51^, Jack CD Lee ^40^, Laura Letchford ^1^ , Nick Levene ^79^, Lisa J Levett ^89^, Michelle M Lister ^56^, Allyson Lloyd ^70^, Joshua Loh ^60^ , Louissa R Macfarlane-Smith ^85^, Nicholas W Machin ^2 , 47^, Mailis Maes ^3^, Samantha McGuigan ^1^, Liz McMinn ^1^, Lamia Mestek-Boukhibar ^41^, Zoltan Molnar ^6^, Lynn Monaghan ^79^, Catrin Moore ^27^, Plamena Naydenova ^3^, Alexandra S Neaverson ^1^, Rachel Nelson ^1^, Marc O Niebel ^21^ , Elaine O'Toole^48^ , Debra Padgett ^64^, Gaurang Patel ^1^ , Brendan AI Payne ^66^, Liam Prestwood ^1^, Veena Raviprakash ^67^, Nicola Reynolds^86^, Alex Richter ^16^, Esther Robinson ^95^, Hazel A Rogers^1^, Aileen Rowan ^96^, Garren Scott ^64^, Divya Shah ^40^, Nicola Sheriff ^67^, Graciela Sluga, Emily Souster^1^, Michael Spencer-Chapman^1^, Sushmita Sridhar ^1, 3^, Tracey Swingler ^53^, Julian Tang^58^, Graham P Taylor^96^, Theocharis Tsoleridis ^55^, Lance Turtle^46^, Sarah Walsh ^57^, Michelle Wantoch ^86^, Joanne Watts ^48^ , Sheila Waugh ^66^, Sam Weeks^41^, Rebecca Williams^31^, Iona Willingham^56^, Emma L Wise ^25, 29, 31^, Victoria Wright ^54^, Sarah Wyllie ^70^, and Jamie Young ^3^.

**Software and analysis tools**

Amy Gaskin^33^, Will Rowe ^15^, and Igor Siveroni ^96^.

**Visualisation:**

Robert Johnson ^96^.

**1** Wellcome Sanger Institute, **2** Public Health England, **3** University of Cambridge, **4** Health Data Research UK, Cambridge, **5** Public Health Agency, Northern Ireland ,**6** Queen's University Belfast **7** Public Health England Colindale, **8** Department of Medicine, University of Cambridge, **9** University of Oxford, **10** Departments of Infectious Diseases and Microbiology, Cambridge University Hospitals NHS Foundation Trust; Cambridge, UK, **11** Division of Virology, Department of Pathology, University of Cambridge, **12** The Francis Crick Institute, **13** Cambridge Institute for Therapeutic Immunology and Infectious Disease, Department of Medicine, **14** Public Health England, Clinical Microbiology and Public Health Laboratory, Cambridge, UK, **15** Institute of Microbiology and Infection, University of Birmingham, **16** University of Birmingham, **17** Queen Elizabeth Hospital, **18** Heartlands Hospital, **19** University of Edinburgh, **20** NHS Lothian, **21** MRC-University of Glasgow Centre for Virus Research, **22** Institute of Biodiversity, Animal Health & Comparative Medicine, University of Glasgow, **23** West of Scotland Specialist Virology Centre, **24** Dept Zoology, University of Oxford, **25** University of Surrey, **26** Wellcome Centre for Human Genetics, Nuffield Department of Medicine, University of Oxford, **27** Big Data Institute, Nuffield Department of Medicine, University of Oxford, **28** Oxford University Hospitals NHS Foundation Trust, **29** Basingstoke Hospital, **30** Centre for Genomic Pathogen Surveillance, University of Oxford, **31** Hampshire Hospitals NHS Foundation Trust, **32** University of Southampton, **33** Public Health Wales NHS Trust, **34** Cardiff University, **35** Betsi Cadwaladr University Health Board, **36** Cardiff and Vale University Health Board, **37** Swansea University, **38** University of Sheffield, **39** Sheffield Teaching Hospitals, **40** Great Ormond Street NHS Foundation Trust, **41** University College London, **42** Oswaldo Cruz Institute, Rio de Janeiro **43** North West London Pathology, **44** Imperial College Healthcare NHS Trust, **45** NIHR Health Protection Research Unit in HCAI and AMR, Imperial College London, **46** University of Liverpool, **47** Manchester University NHS Foundation Trust, **48** Liverpool Clinical Laboratories, **49** University of Exeter, **50** Royal Devon and Exeter NHS Foundation Trust, **51** Quadram Institute Bioscience, University of East Anglia, **52** Norfolk and Norwich University Hospital, **53** University of East Anglia, **54** Deep Seq, School of Life Sciences, Queens Medical Centre, University of Nottingham, **55** Virology, School of Life Sciences, Queens Medical Centre, University of Nottingham, **56** Clinical Microbiology Department, Queens Medical Centre, **57** PathLinks, Northern Lincolnshire & Goole NHS Foundation Trust, **58** Clinical Microbiology, University Hospitals of Leicester NHS Trust, **59** Viapath, **60** Hub for Biotechnology in the Built Environment, Northumbria University, **61** NU-OMICS Northumbria University, **62** Northumbria University, **63** South Tees Hospitals NHS Foundation Trust, **64** North Cumbria Integrated Care NHS Foundation Trust, **65** North Tees and Hartlepool NHS Foundation Trust, **66** Newcastle Hospitals NHS Foundation Trust, **67** County Durham and Darlington NHS Foundation Trust, **68** Centre for Enzyme Innovation, University of Portsmouth, **69** School of Biological Sciences, University of Portsmouth, **70** Portsmouth Hospitals NHS Trust, **71** University of Warwick, **72** University Hospitals Coventry and Warwickshire, **73** Warwick Medical School and Institute of Precision Diagnostics, Pathology, UHCW NHS Trust, **74** Genomics Innovation Unit, Guy's and St. Thomas' NHS Foundation Trust, **75** Centre for Clinical Infection & Diagnostics Research, St. Thomas' Hospital and Kings College London, **76** Department of Infectious Diseases, King's College London, **77** Guy's and St. Thomas’ Hospitals NHS Foundation Trust, **78** Centre for Clinical Infection and Diagnostics Research, Department of Infectious Diseases, Guy's and St Thomas' NHS Foundation Trust, **79** Princess Alexandra Hospital Microbiology Dept. , **80** Cambridge University Hospitals NHS Foundation Trust, **81** East Kent Hospitals University NHS Foundation Trust, **82** University of Kent, **83** Gloucestershire Hospitals NHS Foundation Trust, **84** Department of Microbiology, Kettering General Hospital, **85** National Infection Service, PHE and Leeds Teaching Hospitals Trust, **86** Cambridge Stem Cell Institute, University of Cambridge, **87** Public Health Scotland, 88 Belfast Health & Social Care Trust, **89** Health Services Laboratories, **90** Barking, Havering and Redbridge University Hospitals NHS Trust, **91** Royal Free NHS Trust, **92** Maidstone and Tunbridge Wells NHS Trust, **93** University of Brighton, **94** Kings College London, **95** PHE Heartlands, **96** Imperial College London, **97** Department of Infection Biology, London School of Hygiene and Tropical Medicine.
